# Metabolic programs define dysfunctional immune responses in severe COVID-19 patients

**DOI:** 10.1101/2020.09.10.20186064

**Authors:** Elizabeth A. Thompson, Katherine Cascino, Alvaro A. Ordonez, Weiqiang Zhou, Ajay Vaghasia, Anne Hamacher-Brady, Nathan R. Brady, Im-Hong Sun, Rulin Wang, Avi Z. Rosenberg, Michael Delannoy, Richard Rothman, Katherine Fenstermacher, Lauren Sauer, Kathyrn Shaw-Saliba, Evan M. Bloch, Andrew D. Redd, Aaron AR Tobian, Maureen Horton, Kellie Smith, Andrew Pekosz, Franco R. D’Alessio, Srinivasan Yegnasubramanian, Hongkai Ji, Andrea L. Cox, Jonathan D. Powell

**Affiliations:** Department of Oncology, Johns Hopkins University School of Medicine, Baltimore, MD 21287, USA; Bloomberg∼Kimmel Institute for Cancer Immunotherapy, Johns Hopkins University School of Medicine, Baltimore, MD 21287, USA; Department of Medicine, Johns Hopkins University School of Medicine, Baltimore, MD 21287, USA; Department of Pediatrics, Johns Hopkins University School of Medicine, Baltimore, MD 21287, USA; Department of Biostatistics, Johns Hopkins University Bloomberg School of Public Health, Baltimore, MD 21287, USA; W. Harry Feinstone Department of Molecular Microbiology and Immunology, Johns Hopkins University Bloomberg School of Public Health, Baltimore, MD 21287, USA; Department of Pathology, Johns Hopkins University School of Medicine, Baltimore, MD 21287, USA; Department of Cell Biology, Johns Hopkins University School of Medicine, Baltimore, MD 21287, USA; Department of Emergency Medicine, Johns Hopkins University School of Medicine, Baltimore, MD 21287, USA; Division of Intramural Research, National Institute of Allergy and Infectious Diseases, NIH, Baltimore, MD 21205, USA

## Abstract

It remains unclear why some patients infected with SARS-CoV-2 readily resolve infection while others develop severe disease. To address this question, we employed a novel assay to interrogate immune-metabolic programs of T cells and myeloid cells in severe and recovered COVID-19 patients. Using this approach, we identified a unique population of T cells expressing high H3K27me3 and the mitochondrial membrane protein voltage-dependent anion channel (VDAC), which were expanded in acutely ill COVID-19 patients and distinct from T cells found in patients infected with hepatitis c or influenza and in recovered COVID-19. Increased VDAC was associated with gene programs linked to mitochondrial dysfunction and apoptosis. High-resolution fluorescence and electron microscopy imaging of the cells revealed dysmorphic mitochondria and release of cytochrome *c* into the cytoplasm, indicative of apoptosis activation. The percentage of these cells was markedly increased in elderly patients and correlated with lymphopenia. Importantly, T cell apoptosis could be inhibited *in vitro* by targeting the oligomerization of VDAC or blocking caspase activity. In addition to these T cell findings, we also observed a robust population of Hexokinase II^+^ polymorphonuclear-myeloid derived suppressor cells (PMN-MDSC), exclusively found in the acutely ill COVID-19 patients and not the other viral diseases. Finally, we revealed a unique population of monocytic MDSC (M-MDSC) expressing high levels of carnitine palmitoyltransferase 1a (CPT1a) and VDAC. The metabolic phenotype of these cells was not only highly specific to COVID-19 patients but the presence of these cells was able to distinguish severe from mild disease. Overall, the identification of these novel metabolic phenotypes not only provides insight into the dysfunctional immune response in acutely ill COVID-19 patients but also provide a means to predict and track disease severity as well as an opportunity to design and evaluate novel metabolic therapeutic regimens.

**GRAPHICAL ABSTRACT:** 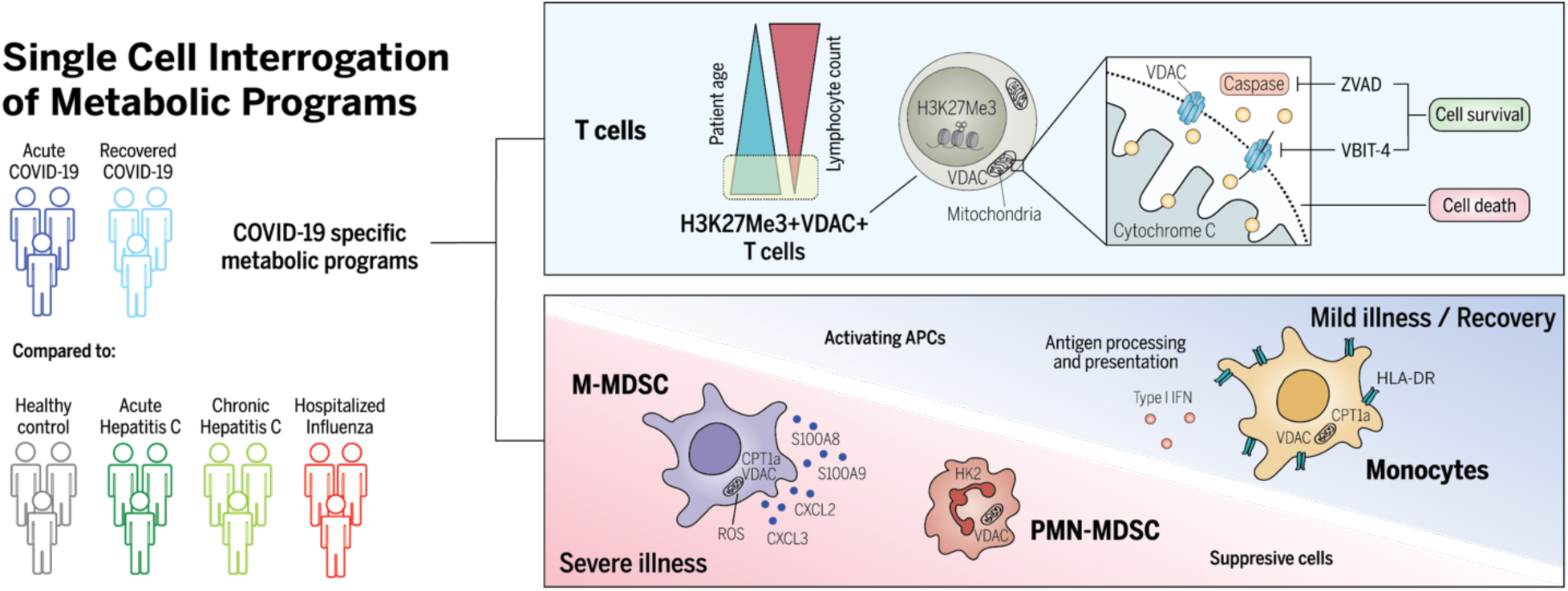

## INTRODUCTION

SARS-CoV-2 is a coronavirus responsible for the COVID-19 pandemic, resulting in over 20 million cases worldwide. While the vast majority of infected patients experience a self-limiting viral syndrome, others develop severe disease leading to pneumonia and acute respiratory distress syndrome (ARDS), accounting for over 1 million deaths globally^1^. At this time, it is unclear why some patients readily resolve infection while others develop severe symptoms. Specifically, it remains to be determined if severe disease is associated with a failure to generate protective immunity, overly robust dysfunctional immune responses, or a combination of both.

To date, severe COVID-19 disease has been associated with multiple changes in peripheral immune profiles including lymphopenia and increased pro-inflammatory cytokines^2^. Recent studies have broadly assessed immune profiles in COVID-19, revealing alterations in both the lymphocyte and myeloid compartments. Preferential loss of CD8+ T cells, increased plasmablasts, neutrophil expansion, decreased pDCs and differential T cell activation have been observed^3-6^. While notable, these observations are indicative of generalized inflammation and thus fail to distinguish specific host deficiencies in SARS-CoV-2 infection versus other viral infections or inflammatory states.

It has become increasingly clear that metabolic reprogramming is not just a consequence of immune activation, but rather plays a critical role in facilitating immune cell differentiation and function^7,8^. For example, effector T cells are characterized by increased expression of molecules necessary to support glycolysis, while memory T cells upregulate expression of molecules involved in oxidative phosphorylation and fatty acid oxidation^9^. Exhausted T cells are characterized not just by the upregulation of inhibitory molecules such as programmed cell death protein 1 (PD-1) and loss of polyfunctionality, but also by mTOR signaling in the absence of productive glycolytic function and anabolic processes^10^. Therefore, we hypothesized that interrogation of immuno-metabolic phenotypes in COVID-19 has the potential to transcend traditional immune cell phenotyping and provide novel insights into distinct functional subsets. To this end, we developed a novel flow cytometry-based proteomic and epigenetic approach that enables the interrogation of metabolic programs at the single cell level (**Table S1, Fig. S1**). Using this approach, we identified disitnct T cell and myeloid subsets in the PBMC of acutely ill COVID-19 patients (COVID-A) that were not found in the PBMC of recovered COVID-19 patients (COVID-R) and patients with hepatitis c and influenza viral infections. Furthermore, these cells provide important mechanistic insight into the immune dysfunction observed in COVID-A patients and the mechanism of pathogenesis.

## RESULTS

### Identification of a Distinct Novel T Cell Subset in the PBMC of Acutely Ill COVID-19 Patients

The high dimensional flow cyotometry-based assay was performed on thawed PBMC from an IRB approved biorepository from patients admitted to the Johns Hopkins Hospital (**Table 1**). We initially focused on T cells within the PBMC given their importance in viral control. Using traditional immunological markers, we observed limited differences in T cell frequencies and phenotype, as previously described^3,11^. Most notably, we detected an increase in the CD4:CD8 ratio and an increase in central memory (Tcm) CD4 T cells in COVID-A patients when compared to healthy controls (**Fig. S2)**.

**Table 1.**
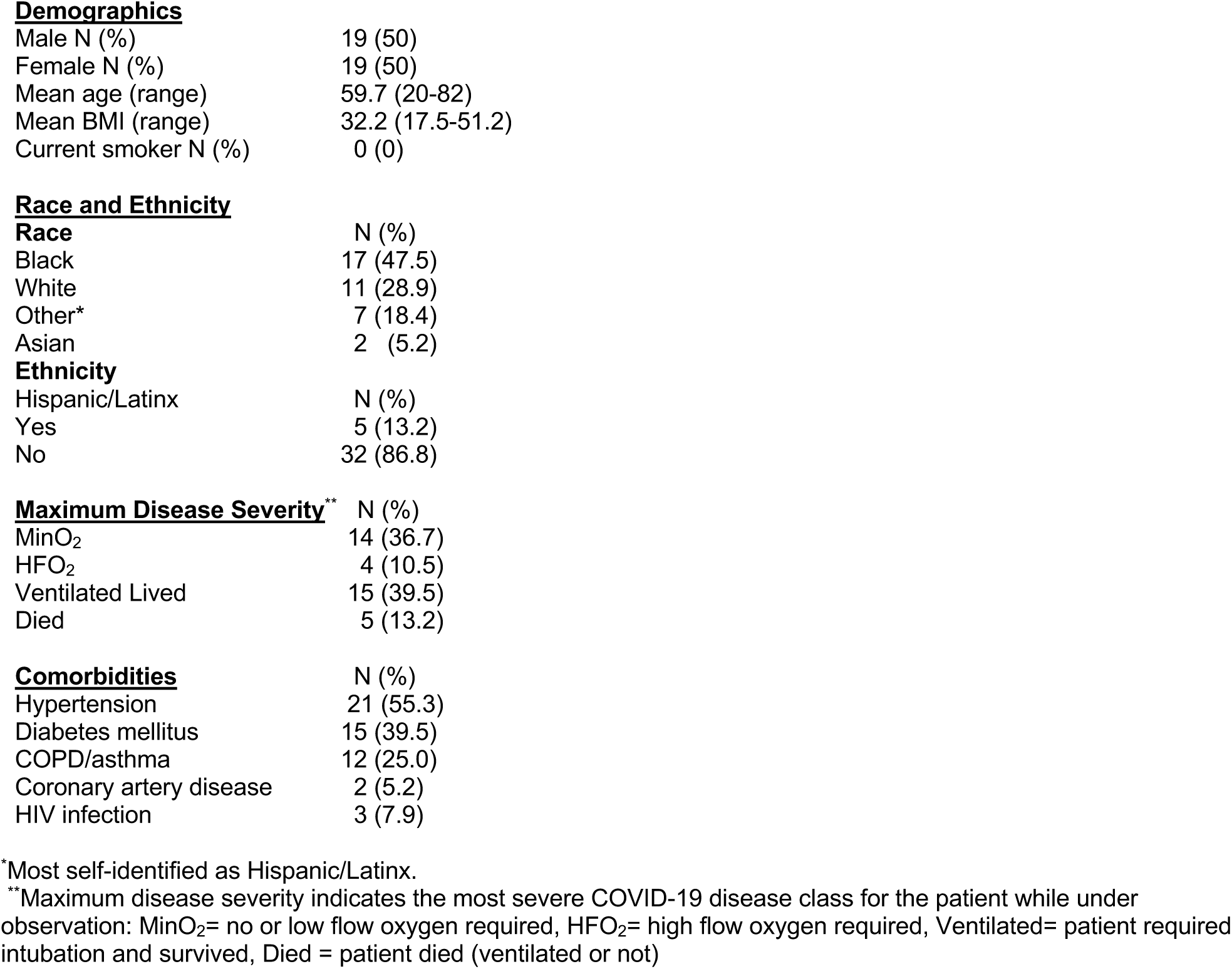
Characteristics of the 38 subjects with acute COVID-19.

**Demographics**
|  |  |
| --- | --- |
| Male N (%) | 19 (50) |
| Female N (%) | 19 (50) |
| Mean age (range) | 59.7 (20-82) |
| Mean BMI (range) | 32.2 (17.5-51.2) |
| Current smoker N (%) | 0 (0) |
| **Race and Ethnicity** |  |
| **Race** | N (%) |
| Black | 17 (47.5) |
| White | 11 (28.9) |
| Other* | 7 (18.4) |
| Asian | 2 (5.2) |
| **Ethnicity** |  |
| Hispanic/Latinx | N (%) |
| Yes | 5 (13.2) |
| No | 32 (86.8) |
| **Maximum Disease Severity\*\*** |  |
| MinO<sub>2</sub> | 14 (36.7) |
| HFO<sub>2</sub> | 4 (10.5) |
| Ventilated Lived | 15 (39.5) |
| Died | 5 (13.2) |
| **Comorbidities** |  |
|  | N (%) |
| Hypertension | 21 (55.3) |
| Diabetes mellitus | 15 (39.5) |
| COPD/asthma | 12 (25.0) |
| Coronary artery disease | 2 (5.2) |
| HIV infection | 3 (7.9) |

\*Most self-identified as Hispanic/Latinx.
\*\*Maximum disease severity indicates the most severe COVID-19 disease class for the patient while under observation: MinO<sub>2</sub>= no or low flow oxygen required, HFO<sub>2</sub>= high flow oxygen required, Ventilated= patient required intubation and survived, Died = patient died (ventilated or not)

In stark contrast, unbiased analysis of T cells employing the combined immune and metabolic markers dramatically segregated T cells from healthy controls and COVID-A (**Fig. 1A**). When compared to healthy controls, we observed no differences in expression of the classical markers of T cell subsets CD45RA, CCR7, or KLRG1 (**Fig. S2F**) or the activation markers CD69, Ki67, PD-1, or HLA-DR (**Fig. S2G**). Likewise, levels of metabolic enzymes involved in glycolysis and fatty acid oxidation did not differ between the two groups (**Fig. S2H**). In contrast, we identified a unique population of T cells found in the COVID-A patients characterized by robust upregulation of Voltage Dependent Anion Channel (VDAC) and the epigenetic mark H3K27me3 (**Fig. 1A-B**). H3K27me3 is regulated in part by a-ketoglutarate-mediated jumonji demethylases^12^, while VDAC is a mitochondrial membrane protein involved in metabolite transport, and has been associated with mitochondrial cell death signaling and lupus-like autoimmunity^13,14^. This phenotype was unique in its exclusive upregulation of mitochondrial proteins, without a concomitant upregulation of glycolytic machinery. Classically, upon T cell activation VDAC and H3K27me3 expression increase along with the glucose transporter Glut1 and Hexokinase II (HKII), both of which support activation-induced glycolysis. This traditional activation signature was seen upon TCR stimulation of healthy PBMCs *in vitro* (**Fig. 1C**). However, the unique population of H3K27me3^hi^VDAC^hi^ T cells in the PBMC of COVID-A patients differ from conventional recently activated T cells because they express relatively low levels of Glut1 and HKII (**Fig. 1D**). The H3K27me3^hi^VDAC^hi^ T cells were found both within the naive and memory T cell compartment (**Fig. 1D**) and did not demonstrate enrichment for specific TCR clones (**Fig. 1E**). Expanded analysis of 55 PBMC samples from 38 COVID-A patients (including sequential samples) revealed that these T cells were markedly upregulated (many exceeding 50% of total T cells) in COVID-A patients and were almost completely absent in healthy controls (**Fig. 1F**). Interestingly, while not present in all of the patients queried, every COVID-A patient 70 years of age and older demonstrated frequencies of the H3K27me3^hi^VDAC^hi^ T cells which were 40% or greater in their PBMC (**Fig. 1G**). Overall, these data reveal a novel, distinct population of H3K27me3^hi^VDAC^hi^ T cells in the PBMC of COVID-A patients. The discordance in activation and metabolic programs suggests that these T cells are dysfunctional.

**Figure 1:**
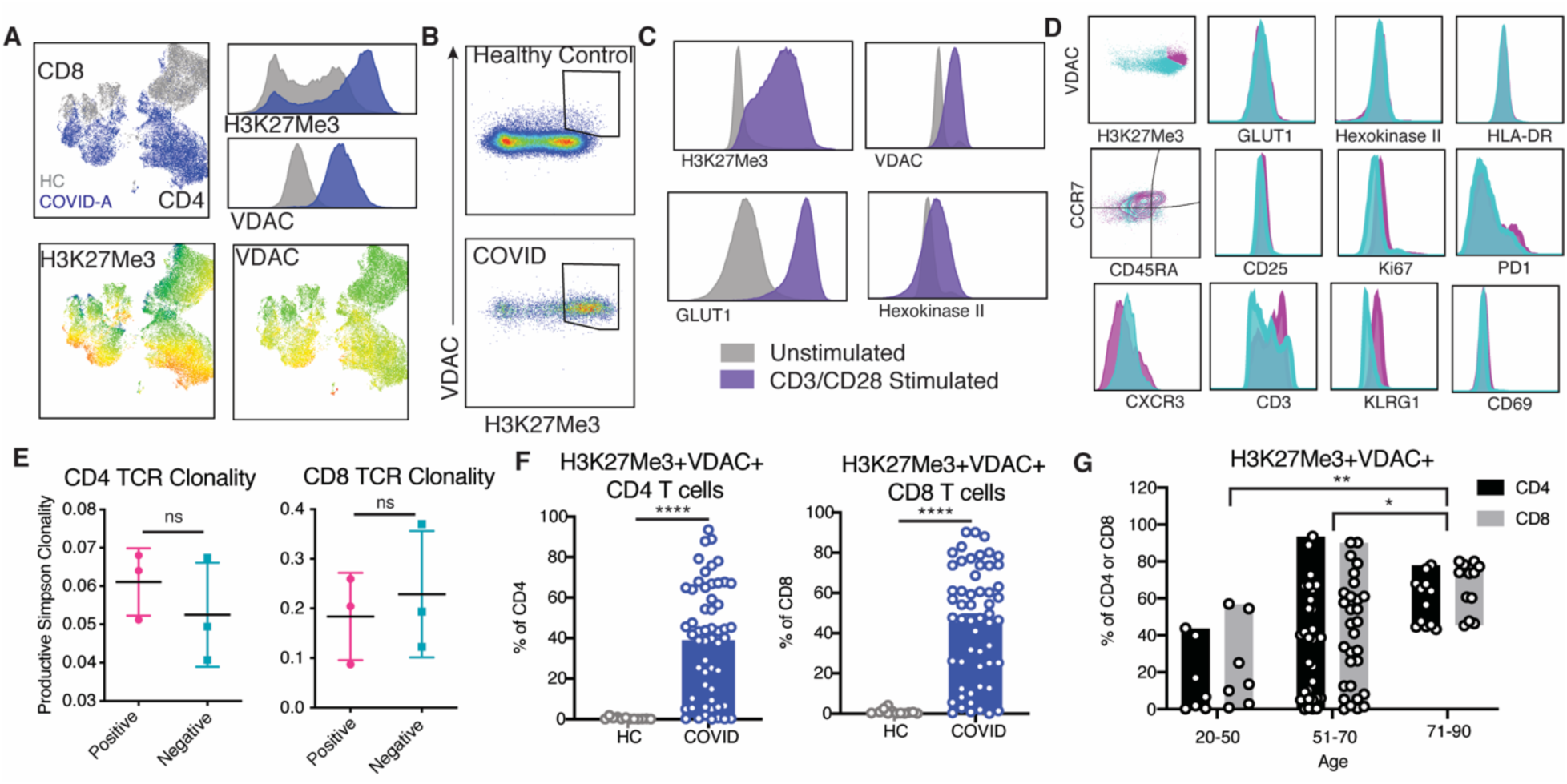
Identification of novel metabolically distinct T cells in COVID-19 patients. A. Concatenated flow cytometry data depicted as UMAP projection of CD3^+^ T cells from healthy control (HC, grey) and hospitalized acute COVID-19 patients (COVID-A, blue). The two markers discovered to drive segregation of the COVID-A and HC cluster, H3K27Me3 and VDAC, are depicted as histogram overlays and MFI heatmap overlays on UMAP projection. B. Representative flow plots of H3K27Me3^+^VDAC^+^ T cells. C. Healthy PBMCs were stimulated for 24 hours with anti-CD3 and anti-CD28 and evaluated for metabolic enzymes using flow cytometry. Representative histograms of unstimulated (grey) and stimulated (purple) cultures. D. Histograms comparing H3K27Me3^+^VDAC^+^ T cells (pink) and remaining T cells (blue) from concatenated pooled COVID-A donors for indicated proteins. E. TCR Simpson Clonality from sorted H3K27Me3^+^VDAC^+^ T cells (pink) and remaining T cells (blue). F. Frequency of H3K27Me3^+^VDAC^+^ as percent of CD4 or CD8 T cells. Each dot represents one individual, significance tested using unpaired Mann-Whitney test. G. Frequency of H3K27Me3^+^VDAC^+^ as percent of CD4 (black) or CD8 (grey) stratified by age of COVID-A patients.

### The Unique Population of H3K27me3^hi^VDAC^hi^ T cells Distinguishes Acutely III COVID-19 Patients From Recovered Patients and Hospitalized Patients Infected With Influenza

By interrogating metabolic programs using our novel assay, we were able to identify a unique population of H3K27me3^hi^VDAC^hi^ T cells in COVID-A patients. We next wanted to determine if this population of T cells was unique to the COVID-A patients. To this end, we interrogated the PBMC of recovered COVID-19 patients (>28 days from diagnosis, COVID-R) and patients with other viral infections to determine if this phenotype was simply a result of ongoing inflammation or general consequence of viral infection. PBMC were obtained and analyzed from patients with acute and chronic viral hepatitis C infection^15^, hospitalized with influenza infection^16^, and those who had recovered from COVID-19 (COVID-R)^17^ (**Table S2**). There appeared to be only subtle differences between these groups in CD4^+^ and CD8^+^ T cell subsets as defined by traditional markers (**Fig. S2**). Yet, when including metabolic markers in the analysis, the COVID-A patients’ T cells again segregated compared to other groups as shown by UMAP projections, indicating a distinct subset of T cells unique to COVID-A patients not accounted for by traditional markers (**Fig. 2A-C**). H3K27me3^hi^VDAC^hi^ T cells were absent in both acute and chronic hepatitis C infection; however, they were present in a subset of both influenza and COVID-R patients (**Fig. 2D-E**). Longitudinal samples were available for eight COVID-A patients, three of which had elevated levels of H3K27me3^hi^VDAC^hi^ T cells at baseline. In these patients, there was a marked reduction in the percentage of these T cells at day 90 during recovery (**Fig S3A-B**).

**Figure 2:**
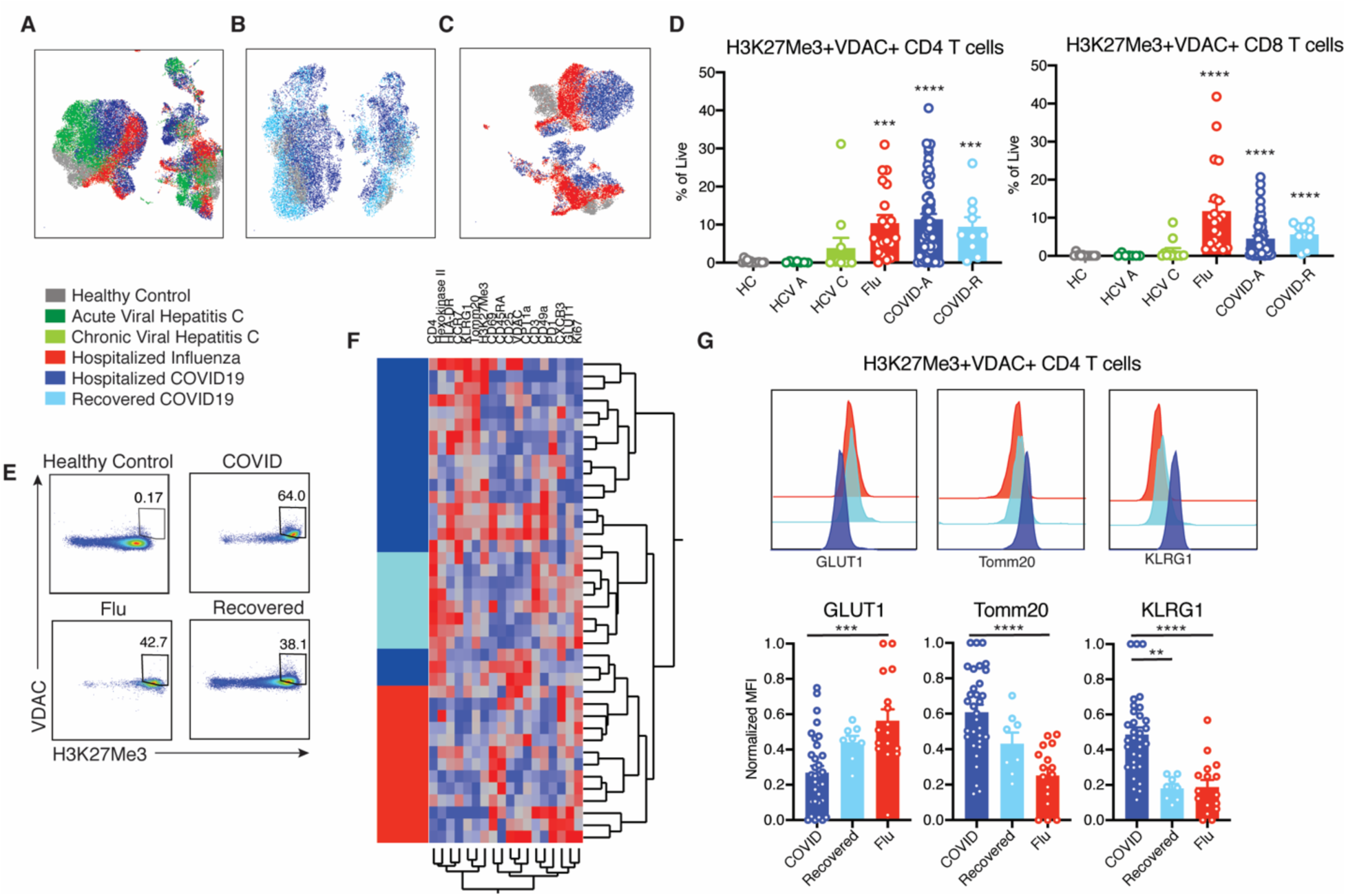
Novel H3K27me3+VDAC+ T cells are unique in COVID-19 compared with other viral infections. A. UMAP projection of pooled donors with active infection, color coded by disease. B. UMAP projection of acute and recovered COVID-19 compared to healthy controls. C. UMAP projection of influenza and acute COVID-19 compared to healthy controls. D. Frequency of H3K27Me3^+^VDAC^+^ CD4 and CD8 T cells as percent of total live cells. E. Representative gating of H3K27Me3^+^VDAC^+^ CD4 T cells from multiple disease states. F. Hierarchical clustering of H3K27Me3^+^VDAC^+^ CD4 T cells based on expression (MFI values) of indicated proteins. Comparison of hospitalized acute COVID-19 infection (dark blue), recovered COVID-19 (light blue) and hospitalized influenza (red). G. Normalized MFI of GLUT1, TOMM20 and KLRG1 in all patients and representative histogram overlays of MFI. Each dot represents one individual, significance tested using unpaired Kruskal-Wallis test compared to healthy control (D) or every combination (G). *p < 0.05, **p < 0.01, ***p < 0.001, and ****p < 0.0001

Importantly, although T cells present in severe influenza infection and during recovery maintained elevated VDAC and H3K27me3 expression, clustering the H3K27me3^hi^VDAC^hi^ CD4 T cells based on overall phenotype showed that these cells robustly clustered based on disease type, indicating qualitative differences in H3K27me3^hi^VDAC^hi^ T cells across disease status (**Fig. 2F**). That is, even though the H3K27me3^hi^VDAC^hi^ T cells were present in the PBMC of the influenza and COVID-R patients, they were distinct from the H3K27me3^hi^VDAC^hi^ T cells in the PBMC of the COVID-A patients. For example, as described earlier, the H3K27me3^hi^VDAC^hi^ T cells from the COVID-A patients demonstrated significantly decreased expression of Glut1, a metabolic marker associated with T cell effector function, compared with COVID-R and influenza (**Fig. 2G**). In contrast, they had significantly increased expression of the mitochondrial protein TOMM20 and KLRG1, a marker associated with T cell senescence and age-related functional defects^18^ (**Fig. 2G**). Thus, despite similar levels of H3K27Me3 and VDAC expression, the metabolic programming of these cells is distinct between acute COVID-19 patients and recovered COVID-19 or hospitalized influenza patients, all of whom recovered. Therefore, these cells represent a population truly unique to acutely ill COVID-19 and may provide insight into the immune dysregulation observed in SARS-CoV-2 infection.

### Dysfunctional Mitochondria in the T Cells of the Acutely III COVID-19 Patients Leads to Apoptosis in a VDAC-Dependent Fashion

In order to elucidate the functional significance of these T cells that were highly prevalent in acute COVID-19, single-cell RNA sequencing was performed on six COVID-A patients, all of which had more than 50% of the T cells of interest as determined by flow cytometry, and three healthy controls. Evaluating gene programs enriched in T cells derived from COVID-A patients revealed an upregulation of cell death programs, as has been reported previously^19^ (**Fig. 3A, red bars**). Furthermore, these data demonstrated evidence of mitochondrial dysfunction, including downregulation of several programs associated with mitochondria function, organization, respiratory chain complex assembly, oxidative phosphorylation, and electron transport coupled proton transport (**Fig. 3A, blue bars**).

**Figure 3:**
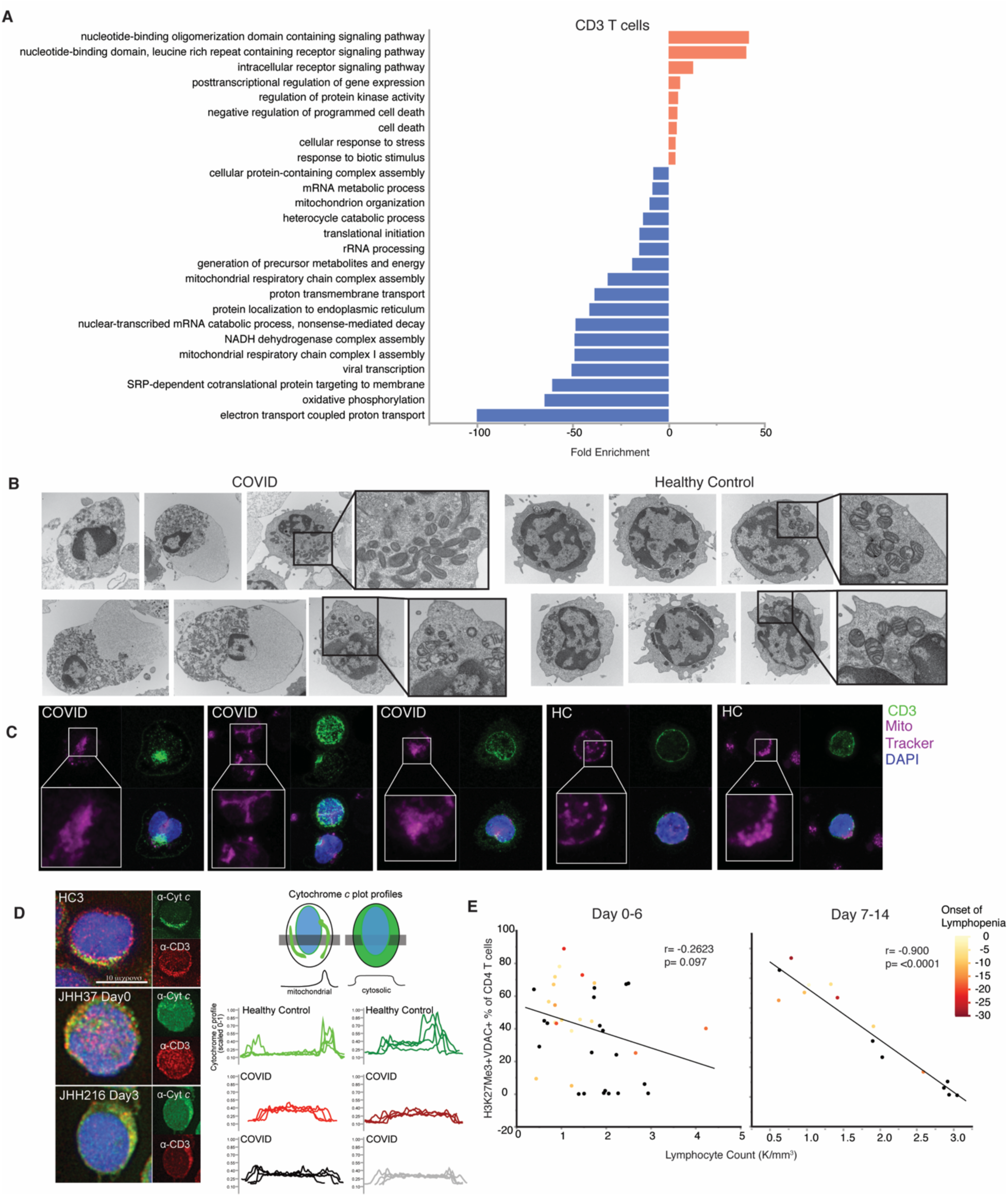
T cells from COVID-19 subjects demonstrate parameters of mitochondrial dysfunction and apoptosis signaling, correlating with development of lymphopenia. A. Single-cell RNA sequencing analysis of 6 COVID-19 subjects and 3 HC were evaluated for CD3+ T cells. Genes distinguishing T cells from COVID-19 patients compared to HC were evaluated for statistical over representation using GO biological processes as gene sets and categorized into higher level annotation using ReviGO. Displayed is the enrichment score for each gene set and color corresponds to programs in upregulated genes (red) and downregulated genes (blue). B. Representative electron microscopy images of PBMCs from a COVID-A patient and a healthy control. C. Representative confocal images of PBMCs from a COVID-A patient and healthy control with mitochondria labeled using MitoTracker Deep Red (pink), CD3^+^ T cells labeled (green) and nuclei labeled with DAPI (blue). D. Representative fluorescence images of PBMCs from 3 COVID-A subjects and one healthy control (left) immunostained for cytochrome *c* (green) and CD3 (red), and nuclei labeled with DAPI (blue). Plot profiles of intracellular cytochrome *c* fluorescence intensity distribution (right). E. Correlation of lymphocyte count with frequency of H3K27Me3+VDAC+ out of total CD4+ T cells from day 0-6 (left) or day 7-14 (right) of study enrollment, including all symptomatic patients. Each dot represents one subject and color corresponds to days since onset of lymphopenia (black = no COVID-19 related lymphopenia developed). Correlation tested using non-parametic Spearman correlation.

To investigate whether elevated levels of VDAC and TOMM20 was indicative of altered mitochondrial function in these T cells, we performed electron microscopy on PBMC from additional COVID-A patients and healthy controls to examine the lymphocyte mitochondria. This analysis revealed markedly dysmorphic, irregularly shaped mitochondria with incomplete cristae in lymphocytes from COVID-A patients compared to lymphocytes from healthy controls, consistent with dysregulated mitochondrial function (**Fig. 3B**). In addition to displaying morphological characteristics of apoptosis, lymphocytes from COVID-A patients showed prominent degenerative changes including large cytoplasmic vacuoles.

We next performed confocal microscopy of PBMC stained with anti-CD3 and MitoTracker Deep Red, which stains mitochondria. CD3^+^ T cells from COVID-A patients demonstrated less distinct mitochondrial staining versus healthy controls (**Fig. 3C**), consistent with the dysmorphic mitochondria observed by EM. Based on these findings and along with the observed decreased mitochondrial programs by RNA-seq and presence of apoptotic cell morphologies by EM, we hypothesized that VDAC might act to facilitate release of mitochondrial cytochrome *c* to the cytosol, leading to apoptosis^20,21^. We therefore performed immunofluorescence analysis of endogenous cytochrome *c* and found it to be present in the cytoplasm of a subset of CD3^+^ T cells from COVID-A patients, whereas in healthy controls, cytochrome *c* was localized to the mitochondria (**Fig. 3D**). Of note, while release of cytochrome *c* into the cytoplasm can lead to rapid cell death, in our procedure, the cells were fixed immediately after thawing, prior to measurement of endogenous cytochrome *c*. Employing this technique enabled us to capture a subset of T cells undergoing active mitochondria-induced apoptosis. Interestingly, the presence of H3K27me3+VDAC+ T cells was highly predictive of the development of lymphopenia during the course of hospitalization. We observed a highly significant (p<0.0001) negative correlation between the frequency of these cells and the peripheral lymphocyte count as measured after 7 days of hospitalization (**Fig. 3E**). This highly significant negative correlation supports the hypothesis that mitochondrial-induced apoptosis is in part contributing to the lymphopenia that develops in the COVID-A patients during the course of their disease.^3,6^. Thus, the unique population of T cells found in the COVID-A patients display mitochondrial dysfunction consistent with cytochrome *c* release into the cytoplasm and subsequent induction of apoptosis. Given these findings and the fact that VDAC can facilitate caspase-mediated cell death^21-23^, we hypothesized that the high expression of VDAC was directly linked to increased susceptibility to cell death in these T cells. To test this hypothesis, we cultured PBMC from COVID-A patients and healthy controls for 48 hours *in vitro* in the presence of media alone, the mTOR inhibitor rapamycin, the VDAC oligomerization inhibitor VBIT-4^13,14^, and the global caspase inhibitor ZVAD (**Fig. 4A**). We observed decreased T cell survival in media alone from the PBMC of COVID-A patients when compared to healthy controls, confirming increased cell death in COVID-A T cells. Interestingly, survival of the COVID-A T cells was rescued with both the VDAC oligomerization inhibitor and the pan-caspase inhibitor, but not the mTOR inhibitor. These findings are consistent with the fact that VDAC oligomerization and interaction with BCL2 family proteins is thought to enable pore formation within the mitochondrial outer membrane, allowing for cytoplasmic release of cytochrome *c*, which initiates the caspase cascade to induce cellular apoptosis^21-23^. Notably, neither VBIT-4 nor ZVAD enhanced survival of the healthy T cells, demonstrating that this mechanism of cell death contributes to loss of viability in the COVID-A T cells but not in the T cells from the healthy controls. Interestingly, the COVID-A T cells treated with ZVAD and VBIT-4 treatment demonstrated robust response to anti-CD3 + anti-CD28 stimulation, similar to the healthy T cells, indicating the ability to functionally rescue the T cells (**Fig. 4B**). These data suggest that VDAC is promoting cell death in these T cells (**Fig. 4C**).

**Figure 4:**
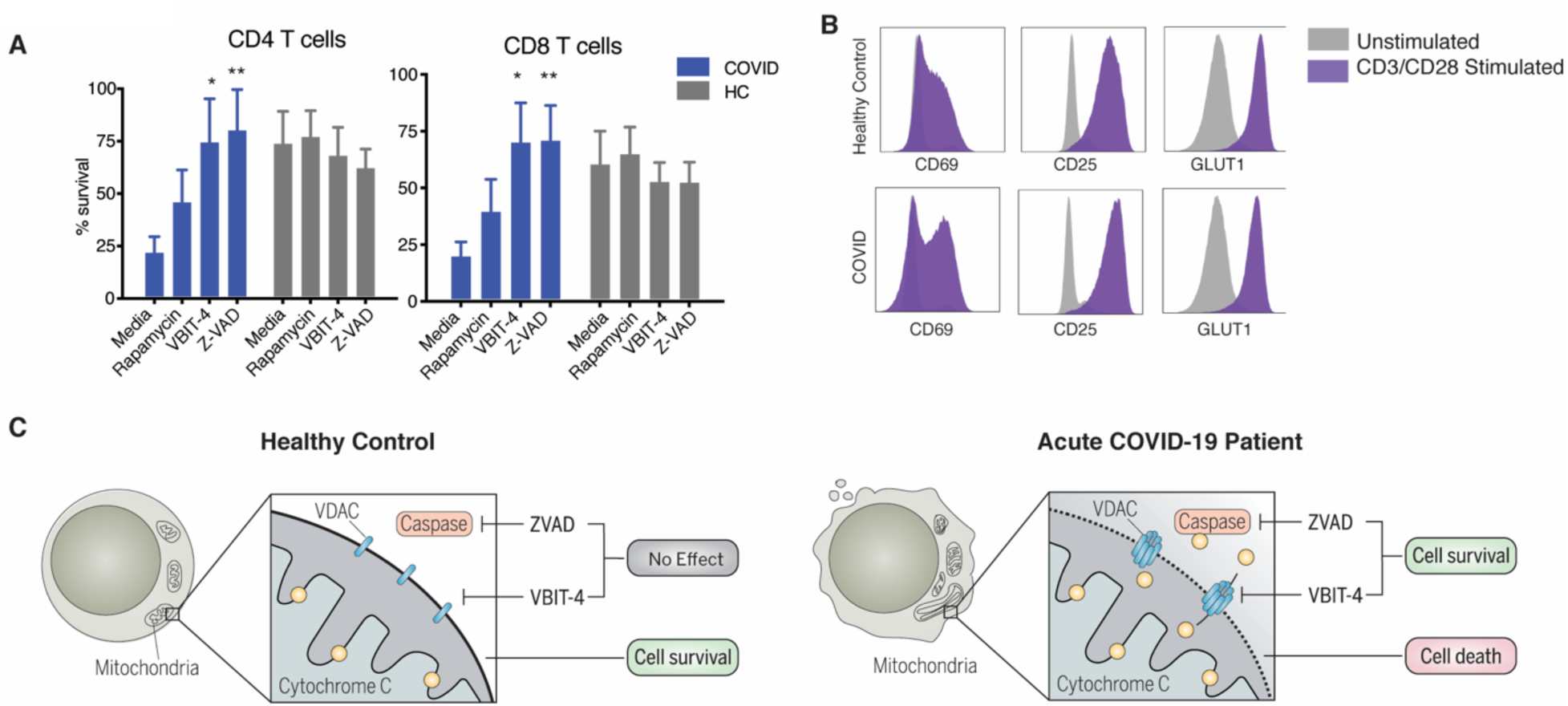
Loss of T cell survival can be rescued by targeting VDAC or caspases. A. PBMCs from acute COVID-19 patients or healthy controls were cultured for 48 hours in media, rapamycin (100nM), VBIT-4 (300nM), or ZVAD (60nM). T cell survival was calculated as the percent CD4 or CD8 T cells remaining from initial plating. Significance tested using two-way ANOVA with each drug compared to the media control, n=9. B. T cells were stimulated with anti-CD3/CD28 (purple) for 48 hours and surviving T cells from COVID-19 patients are able to respond by upregulating HLA-DR, CD69, CD25 and GLUT1 to the same extent as healthy controls compared to unstimulated controls (grey). C. Graphical depiction of proposed mechanism of mitochondrial cell death signaling in COVID-19 T cells. *p < 0.05, **p < 0.01, ***p < 0.001, and ****p < 0.0001

In light of the propensity of the T cells from the COVID-A patients to undergo apoptosis, we wanted to verify that our results did not reflect differences in cell survival after freezing. When we compared fresh and frozen PBMC from four COVID-A patients we observed an increased frequency of the H3K27me3^hi^VDAC^hi^ T cells in the freshly stained PBMC (**Fig. S3C-D**) compared to those frozen and thawed. Thus, our studies are potentially underestimating the frequency of these unique T cells in the COVID-A patients.

### Immune Metabolic Evaluation of B Cells and NK Cells

Using our novel assay we also interrogated the metabolic programs of B cells and NK cells. In contrast to T lymphocytes, we did not observe marked differences in the frequencies of naïve B cells between any viral infections and the healthy controls (**Fig. S4**). Consistent with recent reports, we did observe an increase in the peripheral antibody-secreting cells (ASC) in the COVID-A patients^6^. All of the viral infections were associated with a significant increase in the memory B cells versus healthy controls, while only the COVID-A and hepatitis C patient groups demonstrated an increase in the activated memory B cells. The COVID-A and influenza patient groups both had a higher percent of atypical memory B cells compared to PBMC from the hepatitis C and the COVID-R patient groups. Overall, global UMAP projection demonstrated only subtle phenotypic differences among the different groups in the B cell compartment. When we examined the NK cells, we found no significant differences in frequency when comparing the COVID-A patients, the COVID-R patients and the influenza patients (**Fig. S5**). Conversely, HD flow analysis revealed a population of CD56+ NK cells in the COVID-A patients not present in the influenza patients or in the healthy controls. This population was defined by the upregulation of classical activation markers CD69, Ki67, and CD49a and by increased expression of the metabolic markers TOMM20, CPT1a, and HKII. When we compared NK cells from COVID-A patients with COVID-R patients, we observed differences in CD56+ and CD56bright cells driven mainly by differential expression of markers of activation. While the precise significance of these cells is unclear, their generation can potentially provide important clues into the dysregulated systemic inflammation characteristic of acutely ill COVID-A patients.

### Identification of a Unique Population of VDAC+ Hexokinase II+ Ploymorphonuclear Myeloid Derived Suppressor Cells (PMN-MDSC) in the PBMC of Acutely III COVID-19 Patients but Not Recovered Patients or Patients With Other Viral Illnesses

Next, we examined myeloid cells in the PBMC of the COVID-A patients employing our HD immuno-metabolic assay (**Fig. S6**). Consistent with prior reports, acute COVID-19 was associated with a decreased percentage of myeloid dendritic cells (mDC) and plasmacytoid dendritic cells (pDC) in PBMC and limited differences in monocyte populations^4,6^ (**Fig. S6A-C**).

Visualization of myeloid cells by UMAP projection revealed two distinct populations, which once again only became apparent upon interrogating metabolism (**Fig. 5A-B**). We first identified CD15^+^ granulocytic cells in COVID-A patients, which were entirely absent from the healthy controls (**Fig. 5A-B**). Evaluating the granulocytic cells further revealed a combination of low-density neutrophils, basophils,and polymorphonuclear (PMN)-myeloid derived suppressor cells (MDSC) (**Fig. 3C, Figure S6**). Increased neutrophil counts have been previously observed in COVID-19 patients^4,6^ and low-density neutrophils can be found in the PBMC fraction during inflammation, representing both immature neutrophils and activated/degranulated neutrophils^24^. Immunosuppressive PMN-MDSC are often associated with chronic inflammation, as is seen in cancer, obesity and chronic viral infection^25^. Although low levels of PMN-MDSC were detectable in chronic hepatitis C, there were significantly more in COVID-A patients (**Fig. 3C**). Of note, the PMN-MDSC expressed the highest levels of VDAC within the PBMCs, higher than the H3K27Me3^hi^VDAC^hi^ pre-apoptotic T cells (**Fig. 5D**). However, in contrast to the VDAC+ T cells, the PMN-MDSC had concurrent upregulation of HK2, which is known to associate with VDAC and prevent apoptosis^26^. Thus, the granulocytic suppressor cells (PMN-MDSC) in the PBMC of the COVID-A patients possessed a unique metabolic signature (**Fig. 5E**).

**Figure 5:**
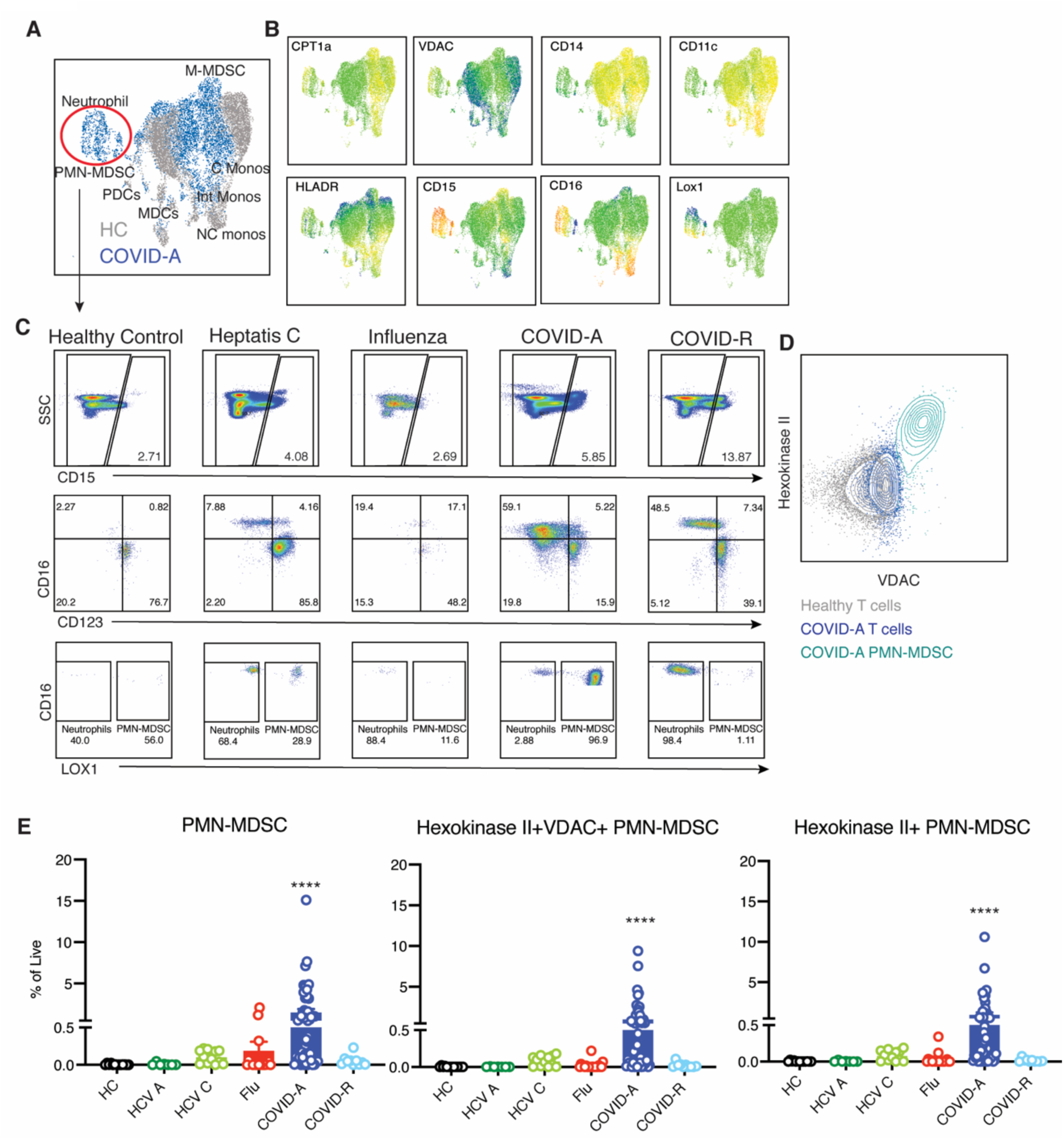
Metabolically distinct granulocytic immunosuppressive myeloid derived suppressor cells in PBMC of COVID-19 patients. A. Concatenated flow cytometry data depicted as UMAP projection of CD3^-^CD19^-^CD56^-^ myeloid cells from healthy control (HC, grey) and acute COVID-19 patients (COVID-A, blue). B. UMAP projection of MFI heatmap overlays of indicated proteins. C. Representative gating of CD15^+^ granulocytic subsets basophils, eosinophils, neutrophils and PMN-MDSC across all disease states studied. D. Flow plot comparing HKII and VDAC expression in HC T cells (grey), COVID-A T cells (dark blue) and COVID-A PMN-MDSC (light blue). E Frequency of indicated cell subset as percent of total live cells. Each dot represents one individual, significance tested using unpaired Kruskal-Wallis test compared to healthy control. *p < 0.05, **p < 0.01, ***p < 0.001, and ****p < 0.0001

### Identification of a Unique Population of CPT1a+ VDAC+ DR-Monocytic-MDSC (M-MDSC) in the PBMC of Acutely Ill COVID-19 Patients That is Associated With Disease Severity

We next identified a population of monocytic cells present in COVID-A patients that expressed high levels of carnitine palmitoyltransferase 1a (CPT1a), an enzyme found within the mitochondrial membrane that is essential for fatty acid oxidation. Additionally, these myeloid cells also expressed high levels of VDAC (**Fig. 6A-B**). Interestingly, CPT1a has been associated with both inflammasome activation and ROS production^27,28^, and a potential role for inflammasome activation in contributing to the pathogenesis of SARS-CoV-2 infection has been noted by others^29^. When comparing the myeloid cells found in COVID-A to COVID-R patients, there was a similar metabolic program driven by high expression of CPT1a and VDAC, however, there was differential expression of HLA-DR (**Fig. 6C**). During acute infection, a large portion of the CPT1a+VDAC+ myeloid cells were HLA-DR-, and thus classified as monocytic MDSC^30^ (M-MDSC) (**Fig. 6C**). A decrease in HLA-DR expression on myeloid cells during COVID-19 infection has been previously noted by other groups as well^5,29,31^. M-MDSC are known potent suppressors of T cell responses, and in line with this role of immunosuppression, they expressed diminished levels of costimulatory molecules such as CD86 compared to their HLA-DR+ counterparts (**Fig. 6D**). Further, they expressed elevated levels of CCR2, indicating recent migration out of the bone marrow (**Fig. 6D**).

**Figure 6:**
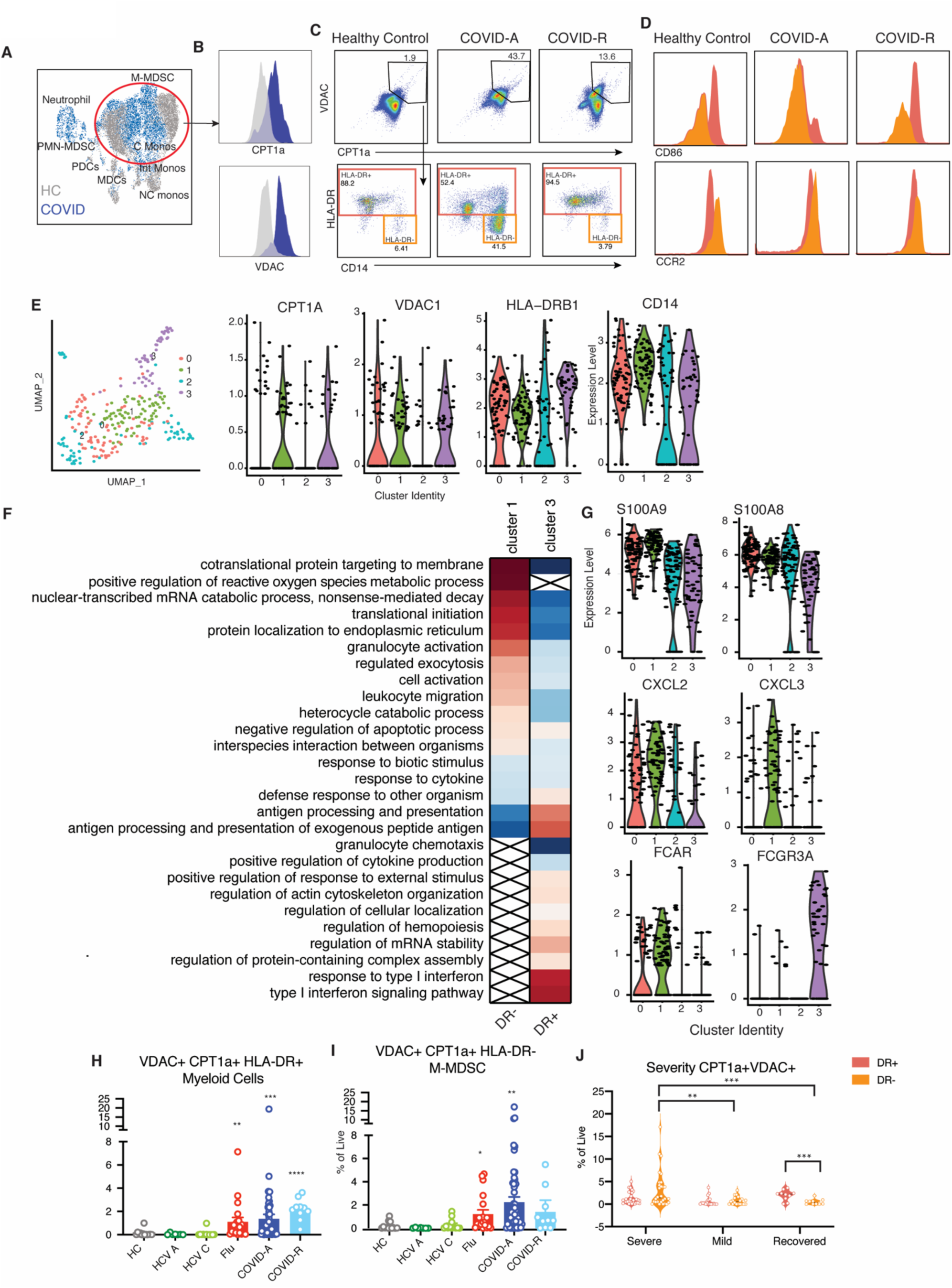
Identification of metabolically distinct monocytic myeloid derived suppressor cells in PBMC of COVID-19 patients track with disease severity. A. Concatenated flow cytometry data depicted as UMAP projection of CD3^-^CD19^-^CD56^-^ myeloid cells from healthy control (HC, grey) and acute COVID-19 patients (COVID-A, blue). B. Histogram overlays of MFI for metabolic markers CPT1a and VDAC from HC (grey) or COVID-A patients (blue). C. Representative gating of CPT1a^+^VDAC^+^ myeloid cells (gated on CD3^-^CD19^-^ CD56^-^ and CD33^+^) and subset of CPT1a^+^VDAC^+^ cells based on HLA-DR expression. D. Histogram overlays of MFI for indicated proteins from CPT1a+VDAC+HLA-DR+ (red) or CPT1a+VDAC+HLA-DR-(orange) cells. E. UMAP projection of scRNA seq of myeloid cells from three COVID patients with detectable CPT1a^+^VDAC^+^ myeloid cells by flow cytometry colored by identified clusters 0-3. Expression of indicated genes within cluster 0-3. Each dot represents a single cell. F. Genes identifying cluster 1 (CPT1a^+^VDAC^+^ HLA-DR^dim^) and cluster 3 (CPT1a^+^VDAC^+^ HLA-DR^high^) were evaluated for statistical over representation using GO biological processes as gene sets and categorized into higher level annotation using ReviGO. Heatmap color corresponds to the enrichment score in upregulated genes (red) and downregulated genes (blue), x indicates a non-significant enrichment. G. Expression of indicated gene within cluster 0-3. Each dot represents a single cell. H-I. Frequency of HLA-DR^-^ CPT1a^+^VDAC^+^ myeloid cells and HLA-DR^+^ CPT1a^+^VDAC^+^ myeloid cells as percent of total live cells. Each dot represents one individual, significance tested using unpaired Kruskal-Wallis test compared to healthy control. J. Frequency of CPT1a^+^VDAC^+^ DR^+/-^ cells in COVID-A patients were stratified by disease severity (Severe= deceased or on ventilator, Mild= hospitalized with low or high flow oxygen, Recovered= post resolution of COVID-19). Each dot represents one individual, significance tested using two-way ANOVA comparing either HLA-DR^+^ vs HLA-DR^-^ within each category or HLA-DR^+^ or HLA-DR^-^ across each category. *p < 0.05, **p < 0.01, ***p < 0.001, and ****p < 0.0001

Conversely, the HLA-DR^+^ component, composed of both CD14^+^ and CD16^+^ monocytes, was elevated during active infection, and remained high during recovery (**Fig. 6C**). HLA-DR^+^ monocytes are capable of producing robust amounts of cytokines and also providing antigen presentation and T cell costimulation, as indicated by the elevated CD86 expression (**Fig. 6D**). The differential expression patterns of these monocytic populations between the infected and recovered patients may represent a shift from immunosuppressive (DR-, MDSC) to a productive immune response (DR+, stimulatory monocytes).

To better understand the function of the two subsets of CPT1a^+^VDAC^+^ HLA-DR^-^ cells, we assessed single-cell RNA sequencing data from the PBMC of three COVID-A patients that had high levels of the CPT1a^+^VDAC^+^ myeloid cells, as determined by flow cytometry (**Fig. 6E**). The analysis revealed four distinct clusters (**Fig. 6E**). Clusters 1 and 3 both had elevated levels of VDAC and CPT1a, but cluster 1 expressed relatively lower levels of HLA-DR compared to cluster 3 (**Fig. 6E**), corresponding to the two populations of cells identified by flow cyotmetry. Cluster 3 (HLA-DR+) exhibited gene programs associated with a productive immune response, such as antigen processing/presentation and a type I IFN response (**Fig. 6F**). In contrast, cluster 1, which demonstrated decreased HLA-DR expression and contained the cells that were more abundant in acute infection, expressed gene programs associated with reactive oxygen species (ROS) production, exocytosis and targeting proteins to the membrane surface (**Fig. 6E**). Therefore, the gene programs supported the hypothesis of an increased immunosuppressive myeloid compartment during acute infection, which transitions into an immunostimulatory profile during resolution. Specifically, the HLA-DR^-^ cluster expressed high levels of genes for secreted alarmins such as S100A9 and S100A8 and chemokines such as CXCL2 and CXCL3^32^ (**Fig. 6G**). Overall, our data demonstrate that the immune activating DR+ cells are present in both the COVID-A and COVID-R patients (**Fig. 6H**). However, the DR-M-MDSC suppressor cells are found in the COVID-A patients but not the recovered COVID patients (**Fig. 6I**).

The dichotomy based on HLA-DR expression of these unique cells and association with productive or inhibitory immune responses prompted us to examine the relationship between the HLA-DR^+^ and HLA-DR^-^ CPT1a^+^VDAC^+^ myeloid cells and disease severity. The CPT1a^+^VDAC^+^ HLA-DR^-^ cells represent less than 0.5% of total PBMCS of COVID-R patients, however, the percentage of these cells is significantly higher in COVID-A patients requiring mechanical ventilation (on average 3.5% of PBMCs) than in COVID-A patients with less severe disease (0.7% of PBMCS) (**Fig. 6J**). While we do not observe these cells in the (non respiratory) hepatitis c infected patients, the DR-cells are present in the influenza patients (**Fig. 6I**). However, the overall percentage of these cells in the influenza patients is lower than that of the COVID-R patients, consistent with the fact that although these patients have been hospitalized they are overall far less ill than the COVID-R patients and all of them recovered from their infections.

### The Unique T cell and Myeloid Subsets in the PBMC of Acutely III COVID-19 Patients Distinguishes Disease Severity

Finally, while our immune-metabolic approach has defined novel T cell and myeloid subsets in the COVID-R patients that provide insight into the the mechanism of immune dysfunction and pathogenesis, we wondered whether the presence of these cells could be exploited to predict and track severity of disease. To this end, we used a random forest model to identify features most important in distinguishing COVID-19 patients from healthy controls, influenza, or between disease severity and recovery (**Fig. 7A-D**). We found that the proportion of H3K27Me3+VDAC+ CD4 T cells was highly predictive when comparing healthy controls to COVID-19 (**Fig. 7A**). Alternatively, when distinguishing between disease severity or between an otherwise similar respiratory viral disease, the innate compartment was highly informative (**Fig. 7B-C**). As previously described, the absence of pDCs was able to distinguish mild from severe COVID-19 (**Fig. 7C**). However, we also found that the HKII+ PMN-MDSC identified were associated with severe disease and could readily discriminate severe COVID from influenza, recovered and healthy controls (**Fig. 7A-B,D**). In addition, CPT1a+VDAC+ myeloid cells (particularly the presence of HLA-DR-M-MDSCs) was associated with more severe disease (**Fig. 7C**). Finally, as prior publications have shown the association between COVID-19 severity and age, sex, and BMI, we further added these variables to our COVID-19 severity analysis^3^. The feature importance analysis confirmed that in our data set sex and BMI are among the most important features in predicting COVID-19 severity. Interestingly, they were less predictive than the percentage of VDAC^+^CPT1a^+^ myeloid cells, pDC, and H3K27Me3^+^VDAC^+^CD4^+^ cells (**Fig. S7B**). Adding the percentage of VDAC^+^CPT1a^+^ myeloid cells, pDC, and H3K27Me3^+^VDAC^+^CD4^+^ improved prediction of COVID-19 severity compared to basic clinical information (**Fig. S7C**), highlighting the important contribution of the immune cell populations identified to disease severity.

**Figure 7:**
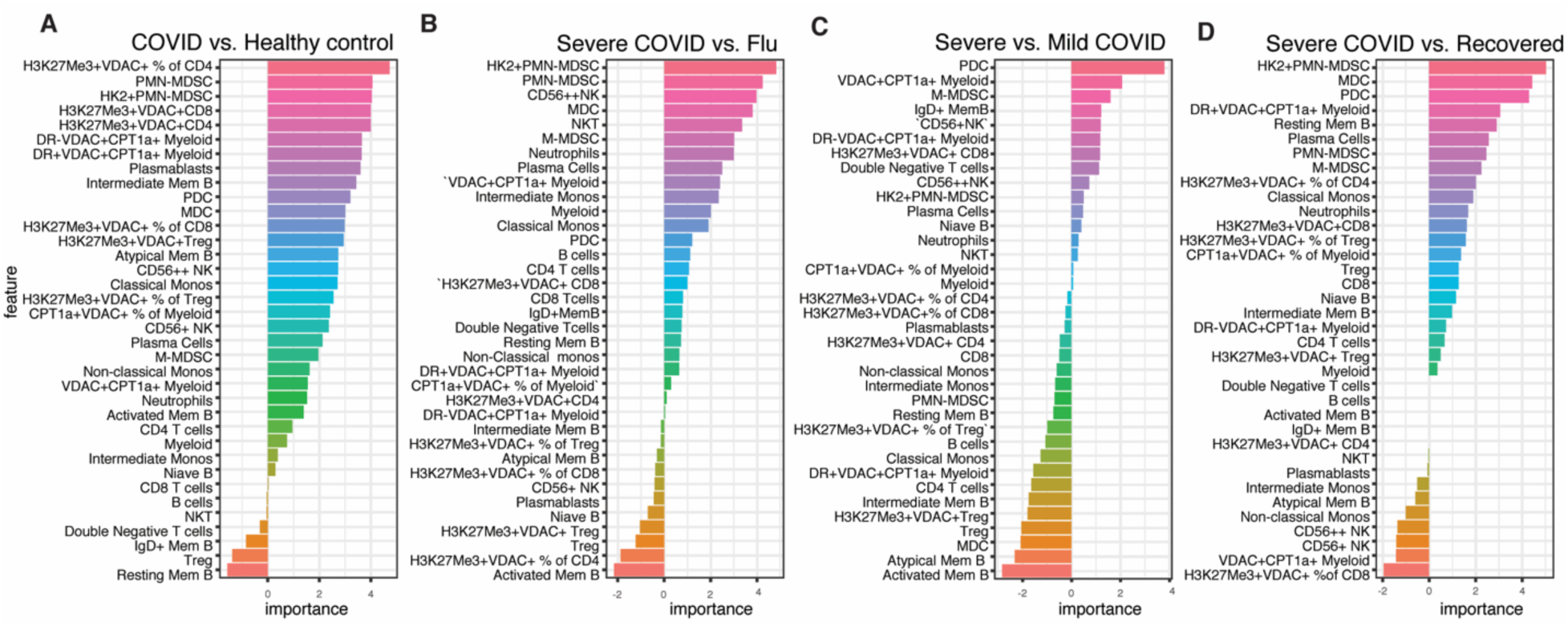
Presence of immune cells with distinct metabolic profiles predicts disease severity. A-D. Feature importance of distinguishing COVID-A patients and healthy controls, COVID-A patients and Flu patients, severe COVID-A patients and COVID-R patients, or severe COVID-A patients and mild COVID-A patients as indicated. Each feature is depicted as a frequency of total live cells, unless otherwise indicated in the case of proportion of cell subset with a specific metabolic phenotype.

## Discussion

Considering that metabolic reprogramming plays an integral role in immune cell differentiation and function, we devised an assay to interrogate metabolic programs at a single cell level. Previous studies examining PBMC from COVID-19 patients have revealed generalized changes in cellular subsets and cytokines consistent with increased inflammation^3-6^. In contrast, by employing our novel approach we were will able to identify robust, distinct T cell and myeloid subsets that were unique to the PBMC of the COVID-A patients. These previously undescribed cells provide important insight into the immune dysfunction and pathogenesis of SARS-CoV-2 infection. Furthermore, as well as defining novel immune cell subsets, our approach reveals a new line of inquiry to understanding immune responses in general in infection, inflammatory diseases, and cancer.

In trying to understand the mechanism of pathogenesis of SARS-CoV-2 infection, there has been much focus on over exuberant immune responses and cytokine storm ^6,29,33^. In contrast, our data has revealed the presence of cells consistent with dysfunctional and suppressive immune responses. T cells play a critical role in both killing virally infected cells and providing help for effective and prolonged anti-viral antibody production^34^. Our data reveals the presence of populations of H3K27me3^hi^VDAC^hi^ T cells in the COVID-A patients. These cells expressed low levels of Glut1, the glucose transporter necessary to support glycolysis during effector responses. Interestingly, we observed parameters indicative of mitochondrial dysfunction and apoptois signaling in these cells, including altered mitochondrial ultrastructure and release of mitochondrial cytochrome *c* to the cytosol. Furthermore, observed loss of cell viability was reversed by treatment with the pan-caspase inhibitor Z-VAD, suggesting caspase dependent apoptosis. Rescue from cell death was likewise achieved by inhibiting VDAC oligomerization, further supporting a mitochondrial mediated cell death program. VDAC is the most prevalent protein in the mitochondrial outer membrane, and acts as the main transporter of nucleotides and metabolites across the mitochondrial membrane^35^. By interacting with pro-apotpotic BCL2 family members, VDAC oligomerization may allow for pores large enough to release of cytochrome *c* into the cytoplasm^21^.

The prevalence of the H3K27me3^hi^VDAC^hi^ T cells increased with age and such cells were observed in all of the patients over 70 years of age in our study. Interestingly, increased age is a known risk factor for severe COVID-19 and aging is associated with a decline in lymphocyte mitochondria fitness and activity^36,37^. Additionally, increased prevalence of the H3K27me3^hi^VDAC^hi^ T cells correlated with lymphopenia in the COVID-A patients. Lymphopenia has previously been described as a prominent feature of COVID-19 and our data suggest that mechanistically this may be due to mitochondrial-induced cell death^2^. To this end, our data support direct T cell death, as opposed to trafficking to inflamed organs such as the lung^38^. Further, this dysregulated T cell immunity could contribute to a lack of or waning protective immunity or impair the functionality of pre-existing cross-reactive T cell immunity^39,40^. Alternatively, it is possible that increased cell death of T cells is driving some of the dysregulated inflammation and autoimmune-like features characteristic of COVID-19. Finally, our data suggest that therapeutically targeting mitochondrial (metabolic) dysfunction might represent a successful strategy for abrogating disease.

The precise inflammatory mediators leading to the generation of the H3K27me3^hi^VDAC^hi^ T cells has yet to be determined. Several lines of evidence indicate that this T cell phenotype is driven by more than the cytokine environment or TCR stimulation alone. Memory T cell subsets are more responsive to secondary cytokine signaling compared to naïve T cells^41^, however, elevated VDAC and H3K27me3 expression was found equally in the naïve and memory compartment. Further, there was no expansion of specific TCR clones within the H3K27me3^hi^VDAC^hi^ phenotype, indicating this is likely not driven by recent antigen-exposure. Because most of the hospitalized COVID-A patients were hypoxic, requiring supplemental oxygen, an interesting possibility is that SARS-CoV-2-induced inflammation in the setting of hypoxia contributes to the generation of these dysfunctional T cells. Indeed, oxidative phosphorylation is required for maintenance of T cell function and is the predominant metabolic pathway utilized by resting T cells^42^. Thus, COVID-19 hypoxia combined with a specific inflammatory environment may lead to this metabolically dysregulated T cell phenotype.

Previous studies have demonstrated increased inflammation and activation of myeloid cells in COVID-19 patients^6,29,31^. However, it still remains unclear which of these responses are protective and which are pathogenic. Our identification of unique myeloid populations in the COVID-A patients shed light on the role of these cells. HKII+ VDAC+ PMN-MDSC were exclusively upregulated in the COVID-A patients, suggesting that this population of cells is either contributing to pathogenesis or the consequence of dysregulated inflammatory responses. On the other hand, CPT1a^+^VDAC^+^DR^+^ monocytic cells were found in both the acute and recovered patients. DR^+^ monocytes have previously been shown to be associated with productive immune responses in other respiratory viral infections^43^ and single-cell RNA sequence analysis revealed increased antigen processing/presentation and type I IFN responses in such cells. In contrast, the CPT1a^+^VDAC^+^DR^-^ M-MDSC were found exclusively in the COVID-A patients and further, the frequency of these cells was positively correlated with severity of disease. As such, the presence of these suppressor cells seems to be indicative of dysregulated inflammation. Finally, CPT1a has been associated with inflammasome activation, which has been observed in COVID-19 patients, and its role in fatty acid oxidation supports modulators of either fatty acid oxidation or inflammasome signaling as potentially novel therapeutic targets to mitigate disease.

Finally, the unique cellular subsets described herein represent potentially potent biomarkers to predict and track severity of disease. The importance of these novel cell populations is highlighted by their ability to robustly contribute to distinguishing severe and mild COVID-19 and acute COVID-19 from other viral infections. Consequently, tracking these unique cells might identify those at highest risk of disease progression and provide important criteria for enrollment into clinical trials as well as provide a surrogate marker for tracking efficacy of new potential treatments. Together, our data demonstrate the utility of broad immuno-metabolic phenotyping to identify novel subsets of immune cells that have the potential to not only provide insight into disease pathogenesis and predict severity of disease but also, importantly, to define novel metabolic targets for treatment.

## Data Availability

All data is available upon request.

## Acknowledgments

We would like to acknowledge the contribution of the Johns Hopkins IVAR team and the JH-EPICS team and assistance for clinical data coordination and retrieval from the Core for Clinical Research Data Acquisition. The specimens from COVID-19 patients utilized for this study were part of the Johns Hopkins Biospecimen Repository, which is based on the contribution of many patients, their families, research teams, and clinicians. We would also like to acknowledge the Electron Microscopy Lab, Department of Pathology, and the Experimental and Computational Genomics Core at the Sidney Kimmel Comprehensive Cancer Center, JHU for help in this study. Likewise, we would like to thank the patients and teams of the Baltimore Before and After Acute Study of Hepatitis (BBAASH), the Johns Hopkins Center of Excellence in Influenza Research and Surveillance (JH CEIRS), Biomedical Advanced Research and Development Authority (BARDA), and the Convalescent Plasma Donor Study.

