## Supplementary Materials for "Metabolic programs define dysfunctional immune responses in severe COVID-19 patients"

### Materials and Methods

#### Study Design and recruitment

Patients diagnosed with COVID-19 by positive SARS-CoV-2 RNA testing through the Johns Hopkins Healthcare System were enrolled in a protocol designed to generate a biospecimen repository linked to clinical data for investigation (Johns Hopkins Medicine (JHM) IRB 00245545) and another for analysis of research questions specific to immunology (JHM IRB 00255162). Subjects identified as SARS-CoV-2 PCR positive consented to study participation and for clinical information to be linked to their study subject identification number. Subjects are categorized by maximum COVID-19 disease severity score using four groups; minimal oxygen required, high flow nasal cannula required, intubation with survival, and death. Samples, including blood for processing into serum, plasma and PBMC, urine, and swabs of the nasopharyngeal, oropharyngeal, and crevicular spaces were obtained as close to admission as feasible (day 0), 3 and 7 days later, weekly after day 7, and after discharge at regular intervals beginning at day 28 following study entry. Demographic information, clinical laboratory test results, ICD-10 coded diagnoses recorded in the patients' records (comorbidities), medication lists, body mass index, and other clinical parameters were linked to data for all subjects in the study. For this study, COVID-19 infected participants on immunosuppressive medications other than inhaled steroids at the time of enrollment were excluded. Following JHM IRB approval, PBMC samples were obtained under informed consent from: HCV infected patients (JHM IRB NA_0004638), hospitalized patients infected with influenza in 2019 (JHM IRB 00091667) and SARS-CoV-2 convalescent plasma donors (JHM IRB 00248402, JHM IRB 00250798) as previously described (*1, 2, 3*) for comparison to hospitalized acutely infected COVID-19 patients in this study.

#### Sample processing and PBMC isolation

Peripheral blood was collected from hospitalized COVID-19 patients upon enrollment in the study at day 0 for isolation of serum, plasma and peripheral blood mononuclear cells (PBMCs). When possible, patients who remained in the hospital were also sampled consecutively at day 3 and day 7 post-enrollment. Blood processing was performed in BSL2+ laboratory conditions as approved following safety assessments. Blood was centrifuged at 400 x *g* for 5 min to separate cells from plasma. Cells were resuspended in RPMI, underlaid with Ficoll and centrifuged at 400 x *g* for 30 min without break at room temperature. The PBMC layer was then washed twice in RPMI and PBMCs were viably cryopreserved in FBS + 10% DMSO for future use.

#### Immuno-metabolic *ex vivo* flow cytometry staining

All flow cytometry antibodies used for phenotypic and metabolic analysis can be found in table S1. PBMCs from hospitalized COVID-19 patients, hospitalized flu patients, COVID-19 convalescent plasma donors (recovered) and healthy controls were used for phenotypic and metabolic assessment. Cryopreserved PBMCs were thawed in RPMI (Gibco) + 50% FBS (Atlanta Biologicals). Cells were washed once in PBS and immediately stained for viability with Biolegend Live/Dead Zombie NIR Fixable Viability Dye and BD Fc Block™ for 10 min at room temperature. Cell surface staining was performed in 100uL of 20% BD Horizon™ Brilliant Stain Buffer + PBS with surface stain antibody cocktail for 20 min at room temperature. Cells were fixed and permeabilized with eBioscience™ FoxP3/Transcription Factor Staining kit 1x Fixation/Permeabilization reagent for 20 min at room temperature. Cells were washed with 1x Permeabilization/Wash buffer. Intracellular staining (ICS) was performed in 100uL 1x Permeabilization/Wash buffer with ICS antibody cocktail for 45 min at room temperature. Cells were washed once with Permeabilization/Wash buffer then resuspended in 1% Paraformaldehyde for acquisition by flow. Samples were run on a 3 laser Cytek Aurora spectral flow cytometer. FCS files were analyzed using Flowjo v10 (10.6.2.) software. Manual gating strategies for both the T cell and B cell/Myeloid panels can be found in Fig S1. High-dimensional unbiased analysis of cell phenotypes was performed using Flojo plugins Downsample v3 and UMAP.

**FACS cell sorting & TCRseq**
PBMCs from three hospitalized COVID-19 patients were stained with the following antibodies to sort on the identified T cell population of interest: Live/Dead Fixable Aqua, CD3 BV786, CD4 BV605, CD8 BV650, H3K27Me3 PE, Tomm20 AF405. PBMCs were thawed as described and immediately filtered through cell strainer capped FACS tubes to avoid excessive cell clumping. Cells were stained for viability with Live/Dead Fixable Aqua and Fc Block for 10 min at room temperature followed by surface staining with CD3, CD4 and CD8 in 20% Brilliant Stain Buffer for 20 min at room temperature. Cells were washed once with PBS. Fixation/permeabilization was performed using ice cold 70% ethanol for 10 min at -20°C. Cells were washed with 2mL PBS + 0.5% BSA + 5mM EDTA and centrifuged at 2000 rpm for 5 min. ICS was performed for markers H2K27me3 and Tomm20 and cells were stained for 45 min at room temperature. Staining reactions were washed once with 2mL PBS + 0.5% BSA + 5mM EDTA and resuspended in 500uL PBS + 0.5% BSA + 5mM EDTA for sorting. $CD4^+$ and $CD8^+$ cells with $H3K27me3^+/Tomm20^+$ or $H3K27me3^-Tomm20^-$ phenotype were sorted by FACS on a Beckman Coulter MyFlo XDP Cell Sorter. Sorted cells were further processed for TCR sequencing. DNA was isolated on sorted populations using QiaAMP micro DNA kit (Qiagen) per the manufacturer's protocol. DNA was incubated overnight in the final column step and eluted in 25uL buffer before quality was assessed via NanoDrop. TCR VbCDR3 sequencing was performed using the deep resolution Immunoseq platform (Adaptive Biotechnologies) (*4*).

**Single-cell RNAseq**
Single cell RNA-seq libraries were prepared from viably frozen PBMCs using the 10X Chromium platform, and 5' DGE library preparation reagents and kits according to the manufacturer's recommended protocols (10X Genomics, Pleasonton, CA). Briefly, viably frozen PBMCs were rapidly thawed at 37°C and were washed twice in DPBS to remove any dead cells and debris. Cells were counted manually with a hemocytometer and re-suspended in 0.04% BSA in DPBS to a final concentration of 1000 cells/uL. Cells and gel beads were loaded on a Chromium Next GEM Chip G to generate single cell emulsions using the 10x Chromium controller instrument with 5' Library Kit v1.1 reagents (PN1000202, PN1000127, PN1000167, PN1000020, PN1000213). Reverse transcription, cDNA amplification, library preparation, and sample index labelling were performed according to manufacturer's protocols. Libraries were sequenced on a NovaSeq 6000 instrument to achieve a target depth of ~50,000 reads per cell. Sequencing data were aligned and pre-processed to generate cell x gene counts matrix for each sample and also aggregate across samples using the cellranger software (v.3.1.0). These data were then imported into Seurat package (v3.1) for subsequent analysis. Data was clustered and visualized using the UMAP method. To analyze T cells, data was subsetted to include only $CD3^+$ cells. The newly subsetted data was then analyzed for differentially expressed genes between COVID-19 and healthy control samples. The gene list was then evaluated for functional enrichment of GO biological processes gene sets using PANTHER (v. 15.0) with Bonferroni correction for multiple hypothesis testing. GO terms were then condensed using ReviGO with a cutoff of 0.4. The fold enrichment determined by PANTHER was visualized. For myeloid cells, total PBMCs were first clustered using UMAP analysis. Clusters containing myeloid cells were subsetted and re-clustered. One cluster was derived predominantly from three COVID-19 patients that had high levels of $CPT1a^+VDAC^+$ myeloid cells as determined by flow cytometry. To better understand the functionality of these cells specifically, the myeloid cells from these three donors were further analyzed, revealing 4 unique clusters. Clusters 1 and 3 were

analyzed for differential gene expression compared to all other myeloid cells due to the high expression of VDAC and CPT1a, matching the flow cytometry data. The gene list was then evaluated for functional enrichment using the statistical over representation with Bonferroni correction using PANTHER (v. 15.0) and GO biological processes as gene sets. GO terms were then condensed using ReviGO with a cutoff of 0.4. The fold enrichment determined by PANTHER was visualized using JMP 14 Pro.

**Electron Microscopy**

For transmission electron microscopy (TEM) PBMCs were thawed as described and washed once with PBS. Cells were chemically fixed as a cell pellet in 3% glutaraldehyde in 0.1M sodium phosphate buffer (pH 7.3) for 24 hours at 4°C, rinsed in 0.1M sodium phosphate buffer, and post-fixed in 1% osmium tetroxide in the same buffer for 1 hour at room temperature. The cells were dehydrated in a graded series of ethanol, transitioned with toluene, followed by infiltration and embedding in epoxy resin EPON 812 (Polysciences,Inc.). Following heat polymerization of the EPON blocks, semi-thin sections of 1000-2000nm thickness were cut and stained with 1% toluidine blue for visualization by light microscopy. Thin sections of selected areas were cut at a thickness of approximately 70-100nm (pale gold interference color) with a diamond knife (DIATOME), placed on 200 mesh copper grids, and dried at 60°C for 10 minutes. To impart electron contrast, the sections were stained with a saturated solution of uranyl acetate for 10 minutes followed by Reynold's lead citrate for 2 minutes. The sections were examined with a transmission electron microscope (JEOL JEM-1400 Plus TEM) using a lanthanum hexaboride cathode (DENKA) operating at an accelerating voltage of 60-80 keV. Images were acquired using an AMT NanoSprint12: 12 Megapixel CMOS TEM Camera (Advanced Microscopy Techniques).

**Immunofluorescence**

For fluorescence detection of cytochrome *c* and CD3, cells were stained as described previously (*5*). Briefly, PBMCs were thawed and immediately placed on a glass microscope slide using a Cytospin 2 centrifuge (Shandon). After centrifugation, the cell monolayer was fixed with 10% formalin and air-dried. Cells were permeabilized with 0.3% Triton X-100 in PBS for 10 min and blocked with 3% BSA for 45 min. Cells were then incubated with primary antibodies against cytochrome *c* (BD Biosciences, cat.no. 556432) and CD3 (Dako, cat.no. A0452) at 4°C overnight. Fluorescent staining was performed for 30 min at room temperature using highly cross-adsorbed Alexa Fluor 488 and 594 secondary antibodies (Invitrogen). Cells were washed three times in PBS after each incubation step. Following, cells were covered with mounting medium containing DAPI nuclear stain (Sigma-Aldrich) and sealed with a coverslip. Imaging was performed with a DeltaVision Elite microscope system (GE Healthcare), equipped with a Scientific CMOS camera (Chip size: 2560 × 2160 pixels), an UltraFast solid-state illumination, a 60x (N.A. 1.42) oil immersion objective and the UltimateFocus module. Single image slices were acquired and deconvolved (Softworx, Applied Precision). Image preparation and analysis was performed using Fiji (http://fiji.sc/Fiji). Intensity profiles were measured using 'Analyze' and 'Plot Profile' on contrast adjusted images. Fluorescence intensities were normalized and plotted in colors corresponding to displayed images.

For fluorescence detection of MitoTracker Deep Red Dye (ThermoFisher, M22426) PBMCs were isolated fresh and labeled with MitoTracker Deep Red Dye for 20 minutes at 37°C in complete media. Cells were then washed with PBS and stained with CD3 FITC (Clone SK7, BD Biosciences). Slides were coated with 50µg/ml Poly-D Lysine (Sigma, P0899) and vigorously washed before addition of cells. Cells on slides were fixed with 4% paraformaldehyde (methanol free, Thermo Scientific, 28906) for 10 min before washing with PBS. Slides were then blocked in 10% goat serum (Gibco, 16210064) followed by staining with goat anti-mouse AF488 IgG

(ThermoFisher, A-11017). Slides were mounted with Slow Fade Diamond anti-fade reagent with DAPI (Invitrogen, S36964). Cells were imaged with an LSM 880-Airyscan confocal microscope equipped with a PlanApochromat 63×/1.4 NA oil-immersion objective (Carl Zeiss). Images were processed with Zen Black software (Carl Zeiss).

#### *In vitro* T cell stimulations

PBMCs were thawed as described above, cells counted, and resuspended to $1x10^6$ cells/mL in complete media (R10; RPMI 1640/heat inactivated 10% FBS). Cells were plated in 96-well U bottom plates in the presence or absence of anti-CD3/28 stimulating antibodies (0.1mg/mL, Miltenyi), and in the presence or absence of Z-VAD-FMK (60nM, Cell Signaling Technology) or VBIT-4 (300nM, Fischer Scientific). The exact number of cells plated were stained directly *ex vivo* to calculate percent survival (number of T cells at day 0/number of T cells at 48 hours). Plates were cultured at 37°C for 48 hours and cells were stained for flow cytometry. Flow staining was performed as described above using the limited panel consisting of CD3 BV786 (BD Biosciences, cat. no. 563800), CD8 BV480 (BD Biosciences, cat. no. 566121), CD4 PE Cy5 (Biolegend, 317412), H3K27me3 PE (CST, cat. no. 40724) and VDAC1 AF532 (Abcam, cat. no. ab14734).

#### Feature importance and prediction analysis

Using the percentage of each cell population as features and the patients as samples, random forests (RF) were trained to classify patients into different groups using R package caret (*6*). The prediction performance was evaluated using the receiver operating characteristic (ROC) curve derived from leave-one-out cross-validation (LOOCV). Within each fold of LOOCV, the optimal model parameter was determined using a nested LOOCV within the training samples. Feature importance analysis was performed based on the RF models trained using all samples. A feature's importance was calculated using the decrease of accuracy after permuting the corresponding feature in out of bag samples in the RF. Features are ordered by their importance in predicting acute COVID-19 vs. Healthy controls, Severe COVID-19 vs. Flu, Severe COVID-19 vs. Recovered, and Severe vs. Mild COVID-19. For predicting COVID-19 severity, basic clinical variables including age, sex, and BMI were also added as features to RF and feature importance analysis was rerun for predicting Severe vs. Mild COVID-19. Based on the feature importance, two prediction models were rebuilt for predicting severity (Severe vs. Mild COVID-19) using the top-five-ranked features (i.e., percentage of $VDAC^+CPT1a^+$ myeloid cells, PDC, and $H3K27Me3^+VDAC^+CD4^+$ cells, sex, and BMI) or the basic clinical information only (i.e., age, sex, and BMI). The two models' performances were compared based on ROC.

#### Statistical analysis

Statistical calculations were performed in GraphPad Prism 8. Data are shown as mean±SEM unless otherwise noted. Comparison between conditions were performed using non-parametric tests as indicated in figure legends. A p value less than 0.05 was considered significant.

**Table S1. Flow cytometry panels and antibodies used.**

T cell Immuno-metabolic Panel

| | MARKER | FLUOROPHORE | VENDOR | CAT # | CLONE |
|---|---|---|---|---|---|
| **SURFACE** | CD3 | BV786 | BD Biosciences | 563800 | SK7 |
| | CD8 | BV480 | BD Biosciences | 566121 | RPA-T8 |
| | CD45RA | BV570 | Biolegend | 304132 | HI100 |
| | CCR7 | BV650 | Biolegend | 353234 | G043H7 |
| | CD25 | BV510 | BD Biosciences | 563352 | M-A251 |
| | HLA-DR | BV750 | Biolegend | 307672 | L243 |
| | CXCR3 | BV605 | Biolegend | 353728 | G025H7 |
| | PD-1 | BV711 | Biolegend | 329928 | EH12.2H7 |
| | CD4 | PE-Cy5 | Biolegend | 317412 | OKT4 |
| | CD69 | PE-Cy5.5 | ThermoFisher Scientific | MHCD6918 | CH/4 |
| | KLRG1 | PE-CF594 | BD Biosciences | 565393 | 2F1 |
| | CD49a | APC | Biolegend | 328314 | TS2/7 |
| | CD19 | APC-Cy7 | Biolegend | 363010 | SJ25C1 |
| | CD56 | APC-Cy7 | Biolegend | 362512 | 5.1H11 |
| **ICS** | FoxP3 | PacBlue | Biolegend | 320116 | 206D |
| | Tomm20 | AF405 | Abcam | ab210047 | EPR15581-54 |
| | VDAC1 | AF532 | Abcam | ab14734 | 20B12AF2 |
| | CPT1a | AF488 | Abcam | ab171449 | 8F6AE9 |
| | Ki67 | PE-Cy7 | Biolegend | 350526 | Ki-67 |
| | H3K27me3 | PE | CST | 40724 | C36B11 |
| | HK2 | AF680 | Abcam | ab228819 | EPR20839 |
| | GLUT1 | AF647 | Abcam | ab195020 | EPR3915 |

B cell/Myeloid Immuno-metabolic Panel

| | MARKER | FLUOROPHORE | VENDOR | CAT # | CLONE |
|---|---|---|---|---|---|
| **SURFACE** | CD14 | BV605 | Biolegend | 301834 | M5E2 |
| | CD16 | BV785 | Biolegend | 302046 | 3G8 |
| | CD33 | BV570 | Biolegend | 303417 | WM53 |
| | CD11c | BV480 | BD Biosciences | 74392 | B-ly6 |
| | HLA-DR | BV750 | Biolegend | 307672 | L243 |
| | CCR2 | BV510 | Biolegend | 357218 | K036C2 |
| | CD40 | PacBlue | Biolegend | 334320 | 5C3 |
| | CD38 | BV711 | BD Biosciences | 563965 | HIT2 |
| | CD86 | BV650 | Biolegend | 305428 | IT2.2 |
| | IgD | BB790 | BD Biosciences | Custom order | IA6-2 |
| | CD27 | PE-CF594 | BD Biosciences | 562297 | M-T271 |
| | CD21 | PE-Cy5 | BD Biosciences | 551064 | B-ly4 |
| | CD138 | PE-Cy5.5 | Biolegend | 356502 | MI15 |
| | CD15 | PE-Cy7 | Biolegend | 301924 | HI98 |
| | LOX1 | PE | Biolegend | 358604 | 15C4 |
| | CD123 | APC | Biolegend | 306012 | 6H6 |
| | CD3 | APC-Cy7 | Biolegend | 344818 | SK7 |
| | CD19 | APC-Cy7 | Biolegend | 363010 | SJ25C1 |
| | CD56 | APC-Cy7 | Biolegend | 362512 | 5.1H11 |
| **ICS** | Tomm20 | AF405 | Abcam | ab210047 | EPR15581-54 |
| | VDAC1 | AF532 | Abcam | ab14734 | 20B12AF2 |
| | CPT1a | AF488 | Abcam | ab171449 | 8F6AE9 |
| | HK2 | AF680 | Abcam | ab228819 | EPR20839 |
| | GLUT1 | AF647 | Abcam | ab195020 | EPR3915 |

**Table S2. Characteristics of study subjects.**

| | COVID-A | Influenza | COVID-R | Acute HCV | Chronic HCV |
|---|---|---|---|---|---|
| **Demographics** | | | | | |
| Male N (%) | 19 (50) | 9 (43) | 6 (60) | 2 (33) | 7 (70) |
| Female N (%) | 19 (50) | 12 (57) | 4 (40) | 4 (67) | 3 (30) |
| Mean age (range) | 59.7 (20-82) | 46.4 (22-89) | 47.8 (18-81) | 25.8 (24-28) | 30.5 (26-35) |

Supplemental Figure 1

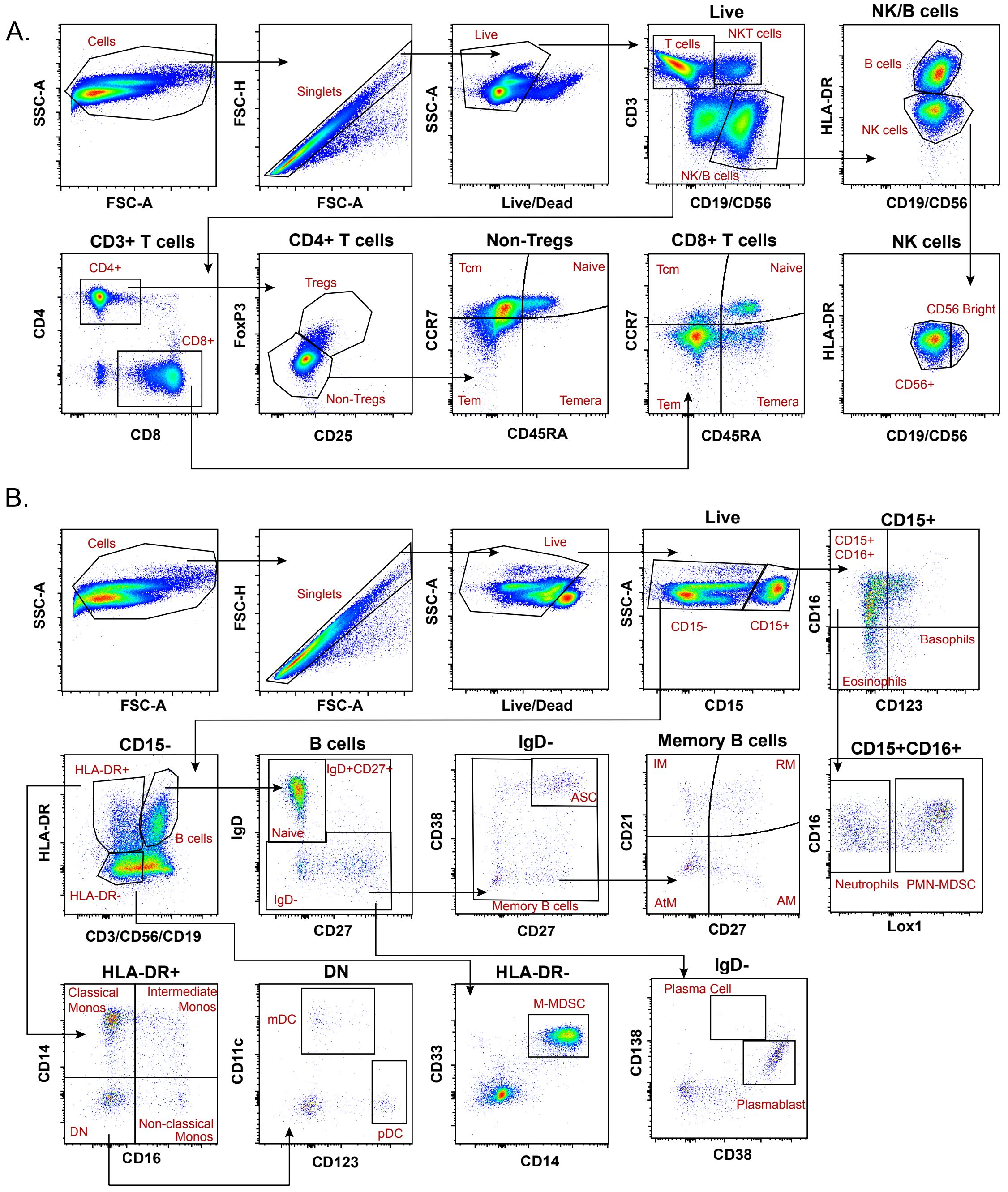


**Fig. S1. Gating strategies for flow cytometry panels.**
Representative flow plots from an acute COVID-19 subject show gating of all peripheral immune cell subsets assessed by one of two immuno-metabolic panels **(A)** T cell panel or **(B)** Myeloid and B cell panel.

Supplemental Figure 2

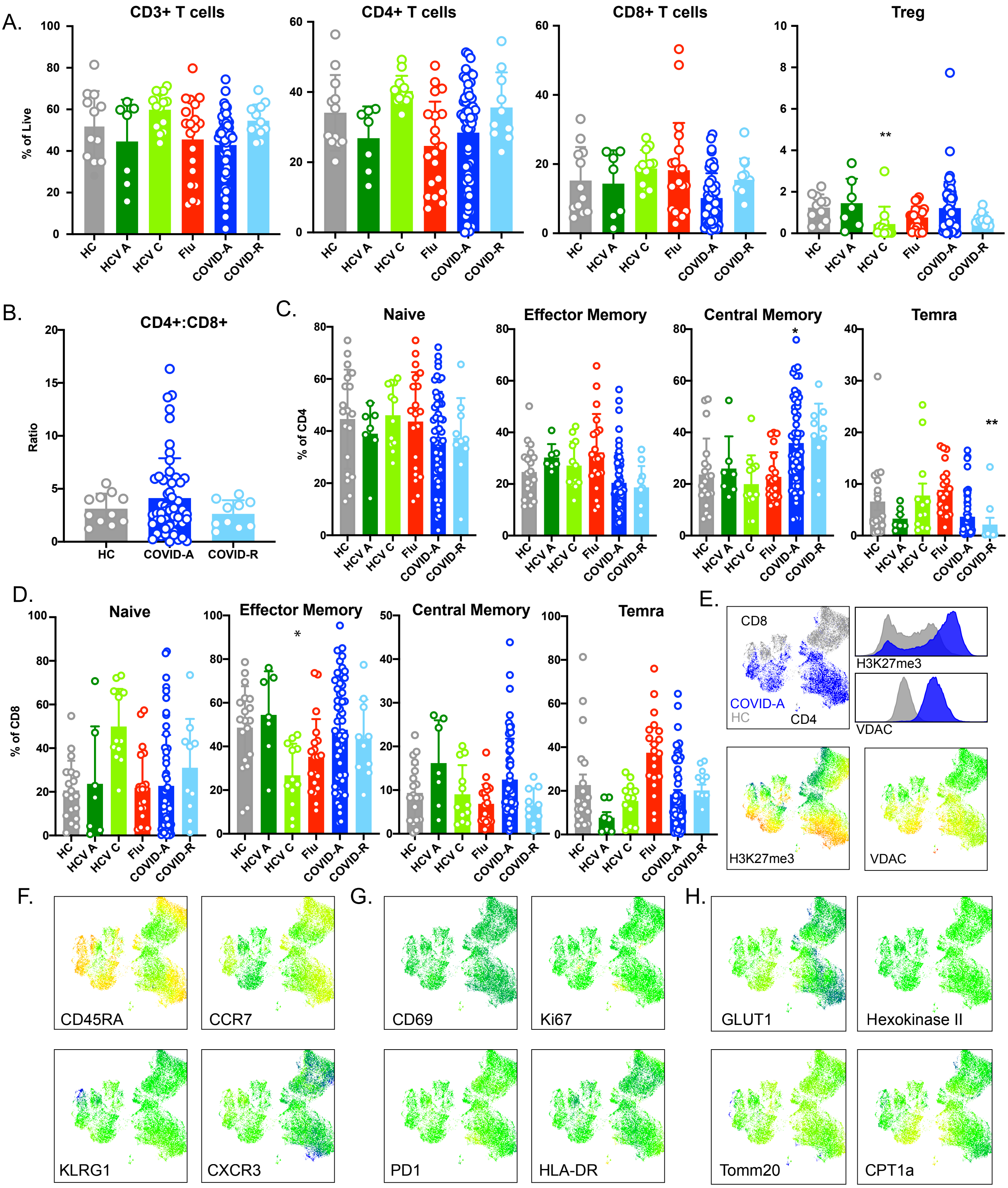


**Fig. S2. Frequencies of T cell subsets and activation markers reveal few COVID19-specific differences.** **(A)** Frequency of indicated cell subset as percent of total live cells. Each dot represents one individual, significance tested using unpaired Kruskal-Wallis test compared to healthy control. **(B)** CD4:CD8 ratio. Each dot represents one individual, significance tested using unpaired Kruskal-Wallis test compared to healthy control. **(C)** Frequency of CD4+ and **(D)** CD8+ T cell subsets shown as percent of CD4 or CD8, respectively. Each dot represents one individual, significance tested using unpaired Kruskal-Wallis test compared to healthy control. **(E)** UMAP projection performed on a subset of COVID-A (blue) and HC subjects (grey). The two markers discovered to drive segregation of the COVID-A and HC cluster, H3K27Me3 and VDAC, are depicted as histogram overlays and MFI heatmap overlays on UMAP projection. **(F-H)** UMAP projection of MFI heatmap overlays of indicated proteins. Signifance is inidicated as compared to healthy control, *p<0.05, **p<0.01, ***p<0.001, ****p<0.0001, if no significance is indicated the test is non-significant.

Supplemental Figure 3

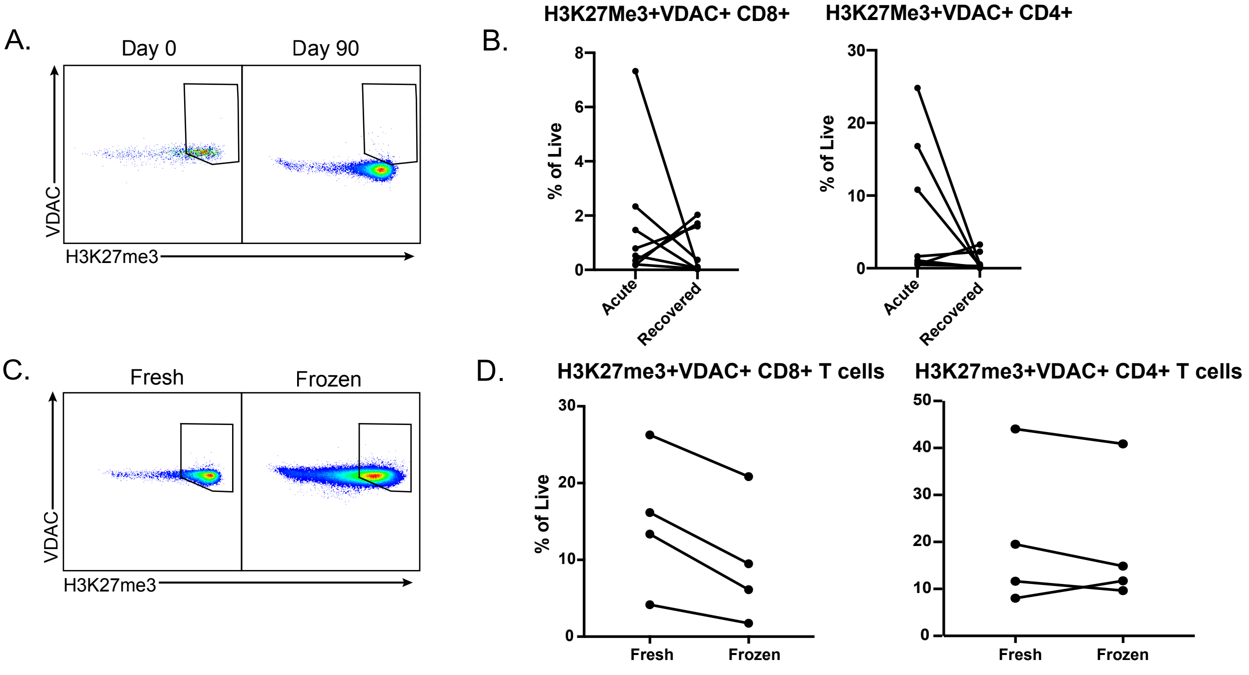


**Fig. S3. H3K27me3+VDAC+ T cell frequencies change over time and after cryopreservation.** **(A)** Representative plots show increased H3K27me3+VDAC+ CD8+ T cells in a COVID-A subject at day 0 enrollment compared to day 90 in the same subject after recovery. **(B)** Frequency of H3K27me3+VDAC+ T cells from 8 total COVID-A subjects with samples available at day 0 enrollment and after recovery for CD8+ T cells (left) and CD4+ T cells (right). Significance tested using Wilcoxon matched-pairs signed rank test. **(C)** Representative plots show increased H3K27me3+VDAC+ CD4+ T cells in a COVID-A subject with cells stained fresh compared to after cryopreservation. **(D)** Frequency of H3K27me3+VDAC+ T cells from 4 total COVID-A subjects tested at day 0 enrollment stained fresh or after cryopreservation for CD8+ (left) and CD4+ (right) T cells. Significance tested using Wilcoxon matched-pairs signed rank test.

Supplemental Figure 4

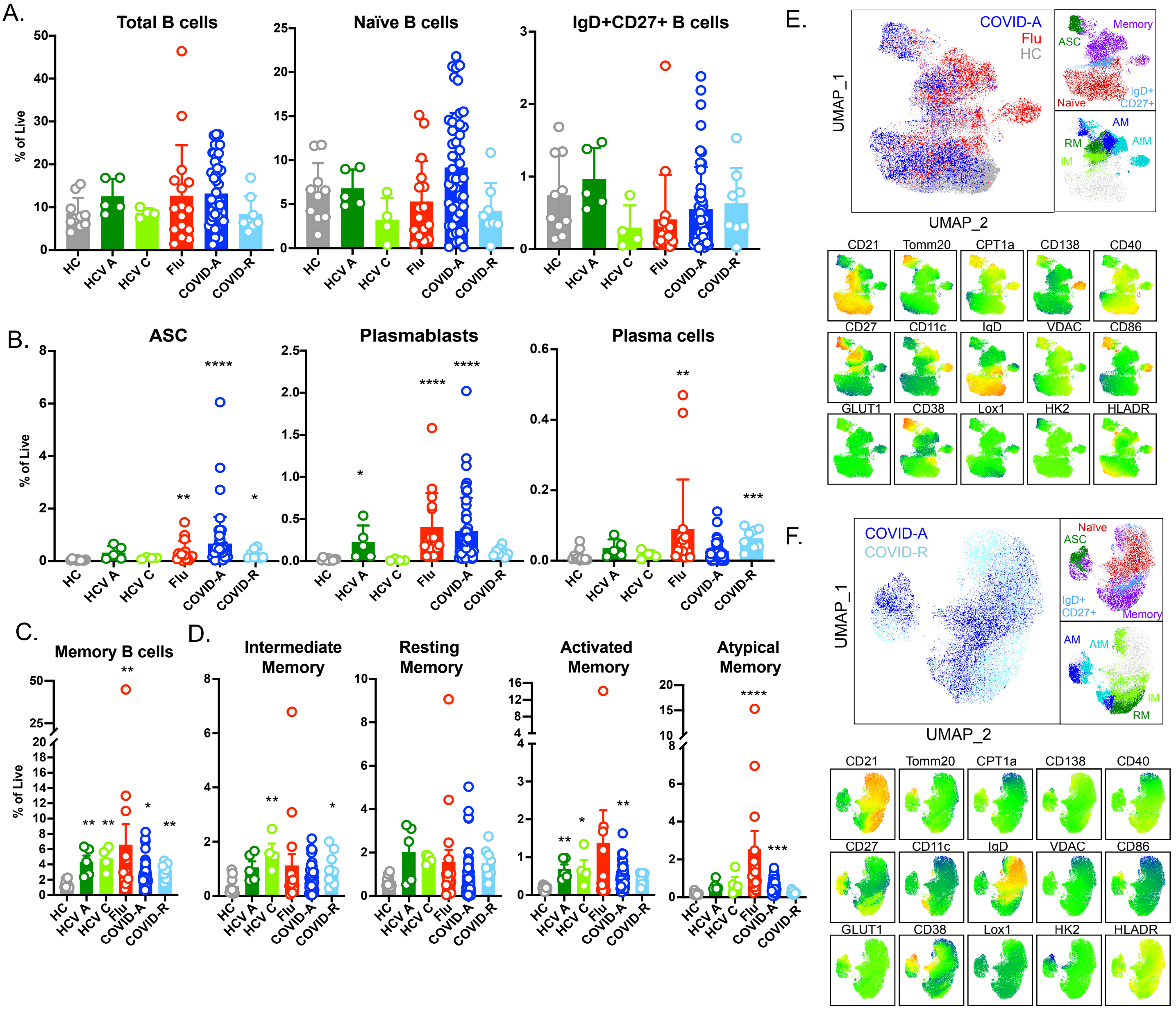


**Fig. S4. B cell frequencies and phenotypes differ in the memory compartment in COVID-19.**
**(A-D)** Frequency of indicated cell subset as percent of total live cells. Each dot represents one individual, significance tested using unpaired Kruskal-Wallis test compared to healthy control. **(E-F)** UMAP projection performed on a subset of COVID-A (blue), hospitalized Flu (red) and HC subjects (grey) **(E)** or COVID-A (blue) and COVID-R (light blue) **(F)**. Manual gating overlays on UMAP projection color code total B cell (top) and memory B cell (bottom) subsets. UMAP projection MFI heat maps of indicated proteins. Signifance is inidicated as compared to healthy control, *p<0.05, **p<0.01, ***p<0.001, ****p<0.0001, if no significance is indicated the test is non-significant.

Supplemental Figure 5

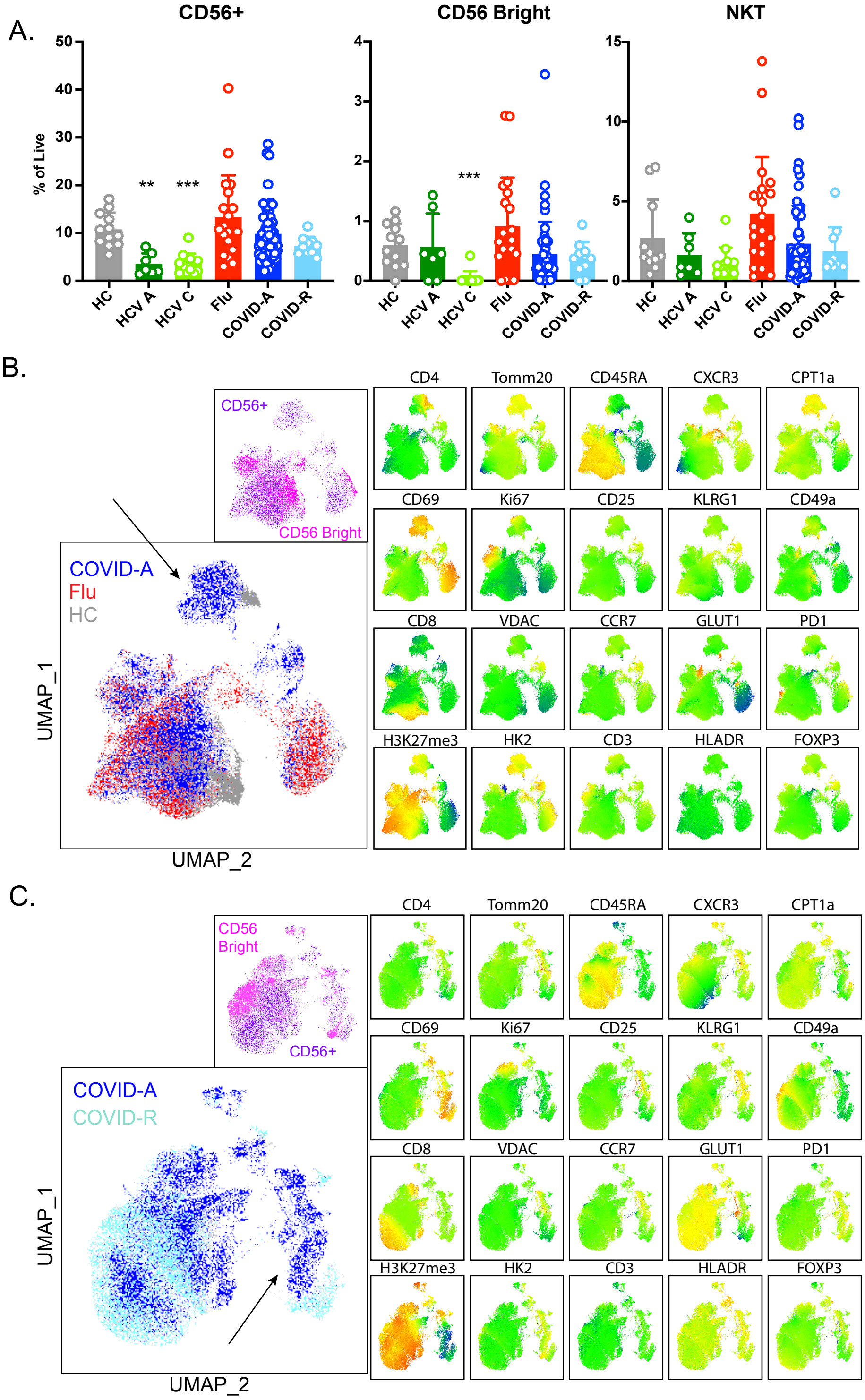


**Fig. S5. Unique NK cell population in COVID-A subjects identified by high dimensional phenotyping analysis.**

**(A)** Frequency of indicated cell subset as percent of total live cells. Each dot represents one individual, significance tested using unpaired Kruskal-Wallis test compared to healthy control. **(B)** UMAP projection of total NK cells performed on a subset of COVID-A (blue), hospitalized Flu (red) and HC subjects (grey) (left). Arrow indicates unique COVID-A specific cluster identified. Manual gating overlays on UMAP projection (top) color code CD56+ (purple) and CD56 bright (pink) cells. UMAP projection MFI heat maps of indicated proteins are shown right. **(C)** Similar analysis as in **(B)** was performed on a subset of COVID-A (blue) compared to COVID-R (light blue) subjects. Signifance is inidicated as compared to healthy control, *p<0.05, **p<0.01, ***p<0.001, ****p<0.0001, if no significance is indicated the test is non-significant.

Supplemental Figure 6

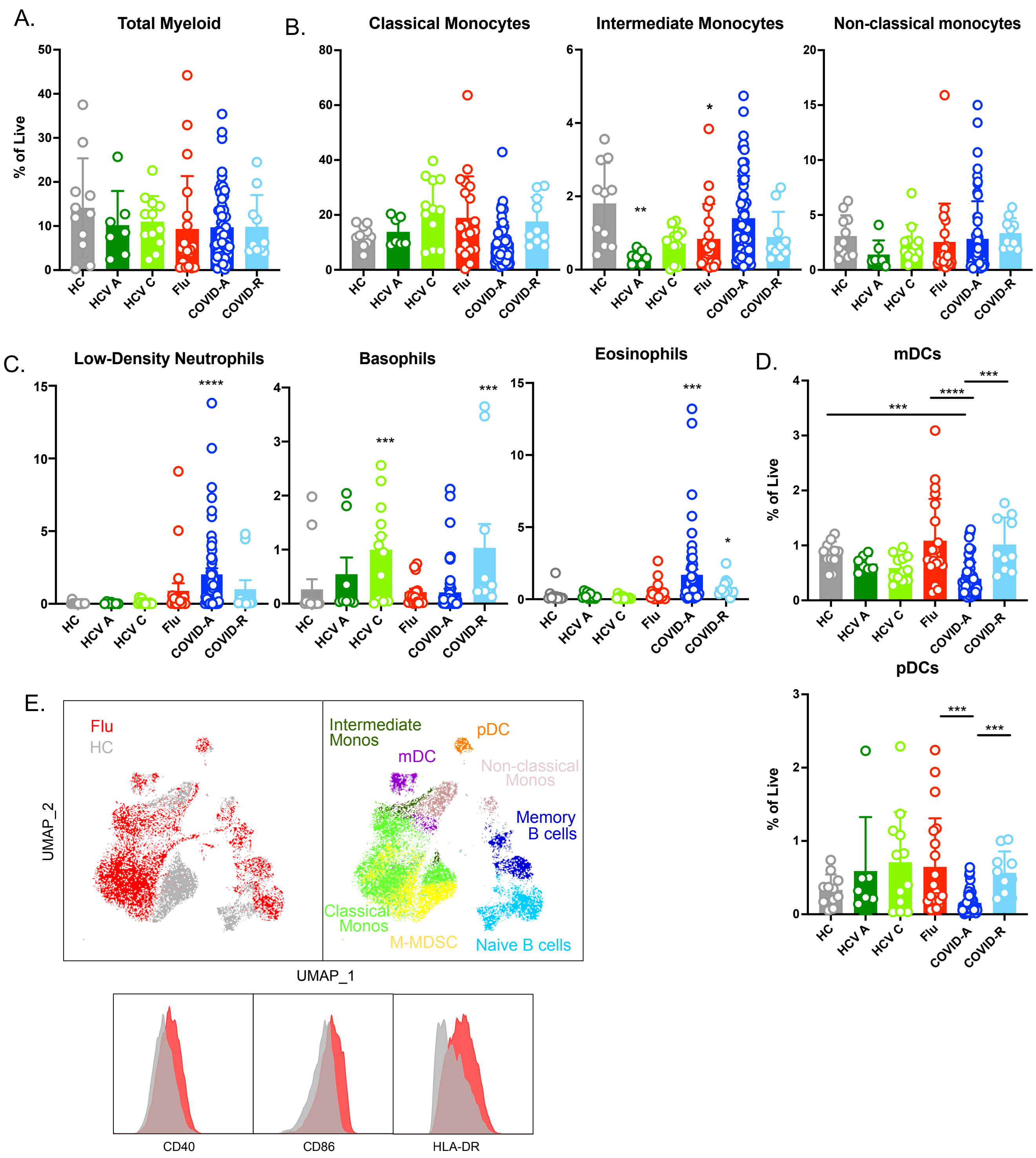


**Fig. S6. Myeloid subsets in viral infections.**
**(A-C)** Frequency of indicated cell subset as percent of total live cells. Each dot represents one individual, significance tested using unpaired Kruskal-Wallis test compared to healthy control. **(D)** Frequency of indicated cell subset as percent of total live cells. To asses how dendritic cell frequencies changed in recovery, significance was tested using unpaired Kruskal-Wallis test comparing all possible combinations. **(E)** UMAP projection of total myeloid cells performed on a subset of hospitalized Flu (red) and HC subjects (grey). Manual gating overlays on UMAP projection color code myeloid and B cell subsets in the UMAP space. MFI histogram overlays of CD14+ myeloid populations of indicated proteins for hospitalized Flu (red) and HC (grey). Signifance is inidicated as compared to healthy control (**A-C**), or between groups (**D**), *p<0.05, **p<0.01, ***p<0.001, ****p<0.0001, if no significance is indicated the test is non-significant.

Supplemental Figure 7

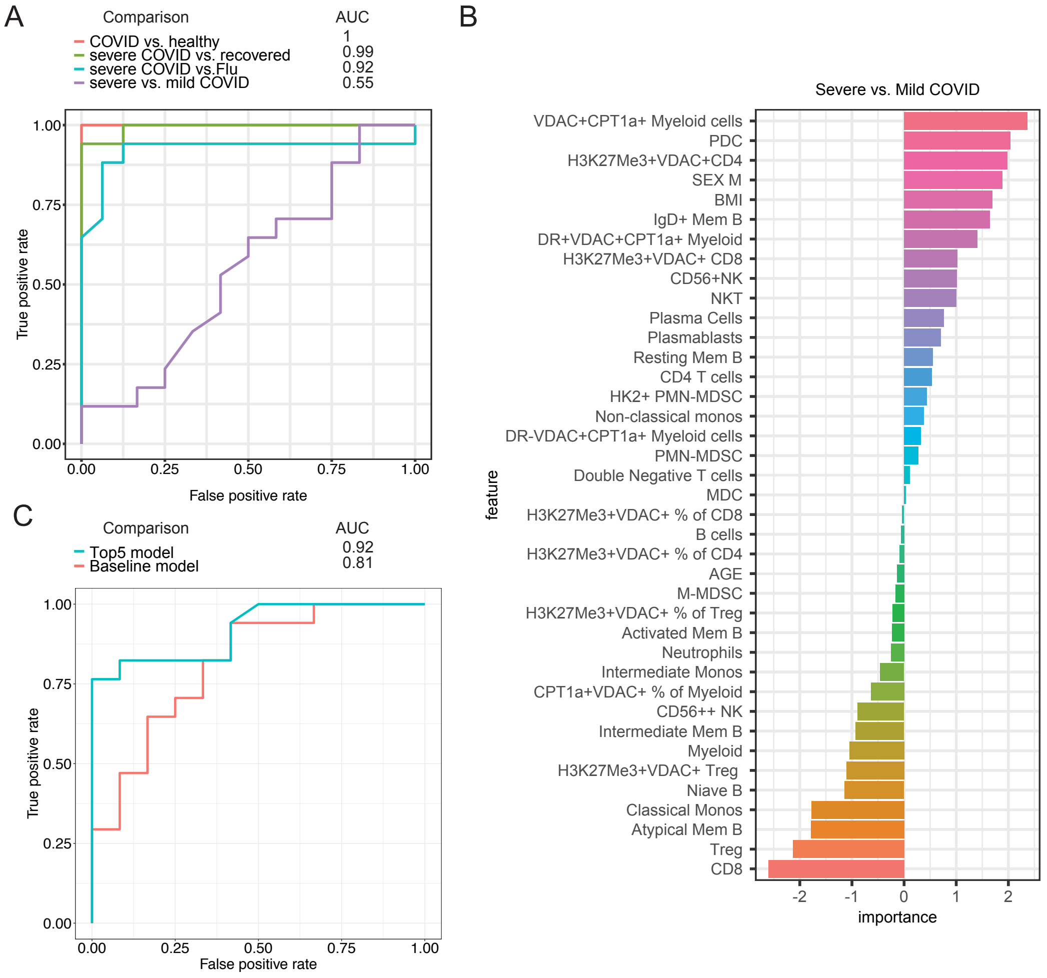


**Fig. S7. Metabolic profile of immune cells predicts disease status.**
**(A)** Receiver operating characteristic (ROC) curves for predicting different groups of patients (i.e., COVID-A vs. Healthy controls, Severe COVID-A vs. COVID-R, Severe COVID-A vs. Flu, and Severe vs. Mild COVID-A). The area under the curve (AUC) is indicated. **(B)** Feature importance analysis after adding basic clinical information (i.e., age, sex, and BMI) to the RF model for classifying severe vs. mild COVID-A. **(C)** ROC curves for comparing the performance of prediction models trained using the top-five-ranked features (i.e., top5) and basic clinical information (i.e., baseline) for classifying severe vs. mild COVID-A. AUC is indicated.
